# Excess death estimates from multiverse analysis in 2009-2021

**DOI:** 10.1101/2022.09.21.22280219

**Authors:** Michael Levitt, Francesco Zonta, John P.A. Ioannidis

## Abstract

Excess death estimates have great value in public health, but they can be sensitive to analytical choices. Here we propose a multiverse analysis approach that considers all possible different time periods for defining the reference baseline and a range of 1 to 4 years for the projected time period for which excess deaths are calculated. We used data from the Human Mortality Database on 33 countries with detailed age-stratified death information on an annual basis during the period 2009-2021. The use of different time periods for reference baseline led to large variability in the absolute magnitude of the exact excess death estimates. However, the relative ranking of different countries compared to others for specific years remained largely unaltered. The relative ranking of different years for the specific country was also largely independent of baseline. Averaging across all possible analyses, distinct time patterns were discerned across different countries. Countries had declines between 2009 and 2019, but the steepness of the decline varied markedly. There were also large differences across countries on whether the COVID-19 pandemic years 2020-2021 resulted in an increase of excess deaths and by how much. Consideration of longer projected time windows resulted in substantial shrinking of the excess deaths in many, but not all countries. Multiverse analysis of excess deaths over long periods of interest can offer a more unbiased approach to understand comparative mortality trends across different countries, the range of uncertainty around estimates, and the nature of observed mortality peaks.

---

Calculation of excess deaths is considered to be a very useful tool for estimating patterns of mortality changes over time in different countries and the impact of major events, such as pandemics (1–3). Excess deaths are meant to capture the composite sum of perturbations in disease incidence and other factors, including social, health care, lifestyle and natural catastrophes that may shape population fatalities in a given year. However, excess death calculations can lead to controversy with different teams of researchers generating markedly different estimates for the same country and year(s) (4–6). The reason is that the calculation of excess deaths requires making analytical choices for which there is no consensus. Specifically, one needs to select a reference baseline period (a time window in the past that will be used for extrapolating how many deaths would be expected in subsequent years) and a projected period (the time window for which an excess death estimate is made by comparing the observed versus expected number of deaths based on the past experience). Moreover, one should decide whether there are any time patterns and what is the form of these time patterns (e.g. whether overall mortality should be declining or increasing over time and, if so, in what form, e.g. linear or spline fit). Empirical work and simulations (4–10) have shown that these choices can make a substantial difference in the obtained excess death estimates.

When results depend on analytical choices, one methodological strategy is to explore the full range of results that can be obtained when a wide range of possible analytical choices and combinations thereof are considered (11–20). Analyses may range from a few dozen to several million different options (e.g. in selecting covariate sets in regressions) (15,17). Different terminology has been used for such approaches that generalize the concept of sensitivity analysis. Commonly used terms are “multiverse analysis” (11–14), “vibration of effects” (16–18) and “multi-analyst analysis” (19,20) (when multiple researchers are each asked to select independently their preferred analysis). Here, we propose a multiverse approach for excess deaths. Instead of making unavoidably arbitrary choices in selecting reference baseline and projected periods, we consider all possible reference baseline periods and projected periods in adjacent year time windows during a lengthy period of interest. Instead of prespecifying time patterns, this multiverse approach allows the data to demonstrate what might be the time patterns and how sensitive the results are to different analytical choices. All possible choices are considered for reference baseline periods (extending as far back as 2009). The multiverse approach also allows us to understand to what extent excess death estimates may shrink when longer projected periods are considered, in the range of 1-4 years. If perturbations lead to excess deaths increases due to the demise of individuals with limited life expectancy (21), then excess death peaks that are seen with short projected periods (e.g. 1 year) will diminish or even disappear when longer projected periods are considered. People who died at some point due to the perturbation would have died very soon anyhow. Conversely, if perturbations result in mortality peaks due to deaths of people who had long life expectancy, extending the projected period window will not have the same impact.

We applied this approach to 33 high-income countries studied before (6) and which have the most reliable data for mortality according to age-stratified groups for the extended period 2009-2021. Our aim here is to propose the multiverse method, illustrate its application, and see how it can offer insights about evolving relative patterns of mortality over many years in each country and how these patterns compare across countries. The multiverse approach focuses on relative comparisons rather than on obtaining absolute estimates of excess deaths during a specific given pandemic period. However, we have also used it to generate absolute estimates of excess deaths during the pandemic period, by considering different types of down weighting of older reference years as opposed to newer reference years.

## RESULTS

### Variability of excess death estimates according to reference baselines

The absolute value of excess death estimates can vary substantially depending on the selection of reference years used for baseline. We considered all 66 possible time windows of whole consecutive calendar years (1 to 11 years long) in the years 2009-2019 as representing baseline values. Table 1 shows the average, standard deviation, minimum, maximum and range for estimates of relative excess deaths (expressed as percentage of expected deaths) for the two-year pandemic period 2020-2021 for each of the 33 countries. The average value is highly correlated with either the maximum or minimum value but not with standard deviation or range (correlation coefficients of 0.96, 0.95, -0.22 and -0.15, respectively). Table 1 also shows the average excess death estimates across 66 possible time windows when different down weighting is applied for older years in the reference range. Figure S1 shows that the average multiverse percentage relative excess death values are highly correlated to the corresponding values calculated with the previously used single reference period of years 2017-19 (6). The multiverse values with the dw3 weighting scheme are very similar to those with the 2017-19 baseline. The multiverse values averaged with equal weights for all 66 baselines are lower by 4.6 percentage points.

**Table 1:** Average, standard deviation, minimum, maximum and range for estimates of relative excess deaths (expressed as percentage of expected deaths referred to as p%) for the two-year pandemic period 2020+2021 for each of the 33 countries.

| Country | Average p% | SD p% | Minimum p% | Maximum p% | Range p% | Average p% dw1 | Average p% dw2 | Average p% dw3 |
| --- | --- | --- | --- | --- | --- | --- | --- | --- |
| Australia | -9.5 | 3.0 | -15.7 | -2.6 | 13.1 | -7.6 | -8.9 | -4.8 |
| Austria | 2.8 | 3.0 | -3.7 | 8.8 | 12.5 | 4.7 | 3.4 | 7.1 |
| Belgium | 0.9 | 3.0 | -5.5 | 8.3 | 13.8 | 2.8 | 1.5 | 5.4 |
| Canada | 0.1 | 2.0 | -6.9 | 4.8 | 11.8 | 1.3 | 0.5 | 2.8 |
| Switzerland | -1.5 | 3.0 | -8.3 | 5.5 | 13.8 | 0.4 | -0.9 | 3.4 |
| Chile | 6.4 | 3.9 | -1.7 | 15.0 | 16.8 | 8.8 | 7.2 | 12.8 |
| Czechia | 10.2 | 3.9 | 1.0 | 18.0 | 16.9 | 12.6 | 11.0 | 15.5 |
| Germany | 1.1 | 1.9 | -4.3 | 4.6 | 8.9 | 2.2 | 1.5 | 3.1 |
| Denmark | -7.6 | 4.0 | -18.6 | -0.2 | 18.3 | -5.1 | -6.8 | -2.9 |
| Spain | 3.6 | 2.2 | -2.6 | 10.9 | 13.5 | 4.9 | 4.0 | 7.1 |
| Estonia | 0.8 | 4.8 | -11.6 | 10.1 | 21.7 | 3.8 | 1.8 | 7.1 |
| Europe | 2.5 | 2.2 | -3.5 | 7.8 | 11.3 | 3.8 | 2.9 | 5.6 |
| Finland | -5.3 | 3.1 | -11.8 | 1.6 | 13.4 | -3.3 | -4.6 | -0.9 |
| France | 2.6 | 2.0 | -3.6 | 6.4 | 10.0 | 3.8 | 3.0 | 5.0 |
| United Kingdom | 4.2 | 1.9 | -1.2 | 10.1 | 11.3 | 5.3 | 4.5 | 7.1 |
| Greece | 5.6 | 2.8 | -1.3 | 10.7 | 12.0 | 7.2 | 6.2 | 8.4 |
| Croatia | 7.0 | 3.1 | -1.2 | 14.9 | 16.1 | 8.9 | 7.7 | 11.5 |
| Hungary | 6.8 | 2.7 | 0.5 | 13.1 | 12.6 | 8.5 | 7.4 | 10.5 |
| Iceland | -7.3 | 2.1 | -12.2 | -2.1 | 10.1 | -6.4 | -7.0 | -4.4 |
| Israel | -1.5 | 2.9 | -7.0 | 4.6 | 11.6 | 0.3 | -0.9 | 2.7 |
| Italy | 5.5 | 2.4 | -0.4 | 10.8 | 11.2 | 6.9 | 5.9 | 8.9 |
| South Korea | -13.8 | 5.3 | -24.7 | -1.2 | 23.5 | -10.4 | -12.7 | -5.8 |
| Lithuania | 8.6 | 3.3 | 2.1 | 18.8 | 16.8 | 10.6 | 9.3 | 14.4 |
| Luxembourg | -2.8 | 3.9 | -10.7 | 3.8 | 14.5 | -0.5 | -2.0 | 1.4 |
| Latvia | 7.1 | 3.2 | -1.0 | 14.0 | 15.0 | 9.0 | 7.7 | 11.1 |
| Netherlands | 2.5 | 2.0 | -2.5 | 7.8 | 10.4 | 3.7 | 2.9 | 5.4 |
| Norway | -9.4 | 3.6 | -16.0 | -1.4 | 14.7 | -7.1 | -8.6 | -3.9 |
| New Zealand | -9.1 | 2.5 | -15.5 | -4.2 | 11.3 | -7.6 | -8.6 | -6.1 |
| Poland | 14.3 | 3.5 | 4.0 | 19.9 | 15.9 | 16.4 | 15.0 | 17.9 |
| Portugal | 2.8 | 2.7 | -4.4 | 8.1 | 12.5 | 4.5 | 3.4 | 6.2 |
| Slovakia | 9.9 | 4.4 | 0.3 | 20.3 | 20.0 | 12.7 | 10.8 | 16.4 |
| Slovenia | 4.7 | 3.4 | -4.0 | 11.8 | 15.7 | 6.8 | 5.4 | 9.4 |
| Sweden | -6.7 | 3.4 | -12.4 | 4.2 | 16.7 | -4.5 | -6.0 | -0.7 |
| United States | 16.7 | 0.8 | 14.4 | 18.7 | 4.3 | 17.1 | 16.8 | 17.7 |

dw: down weighting older years; see Methods for the description of the three different down weighting schemes.

### Stability of relative ranking for the pandemic years’ excess deaths across 33 countries

The estimates of relative excess deaths (as percentage of expected deaths) can be used to compare different countries in a given time period. Despite large variabilities in the absolute estimates, the relative ranking of the 33 countries for a given period of interest was largely unperturbed, regardless of what reference baseline years were chosen. Figure 1 shows the ranking of relative excess death estimates (as percentage of expected deaths) for the pandemic years 2020-2021 in all 66 analyses with different reference baseline windows. The USA had the highest estimates of relative excess deaths among all 33 countries in 50 of 66 analyses, the second highest in 15 analyses and the fourth highest in 1 analysis. Conversely, South Korea had the lowest estimates in 59 of 66 analyses, the second to lowest in 5, the third to lowest in 1, and the sixth to lowest in 1. Eastern European and Balkan countries closely followed the USA in the top excess death ranks consistently. Scandinavian countries, Australia, and New Zealand consistently were placed among the lowest excess death ranks next to South Korea. Other Western European countries typically occupied middle ranks. Figures S2A, S2B and S2C show that the distribution of country ranks for projected periods of 1 year, 3 years and 4 years are similar to that shown in Figure 1 for 2020-2021; summing over more years does blur the ranking of middle-ranked countries.

**Figure 1:**
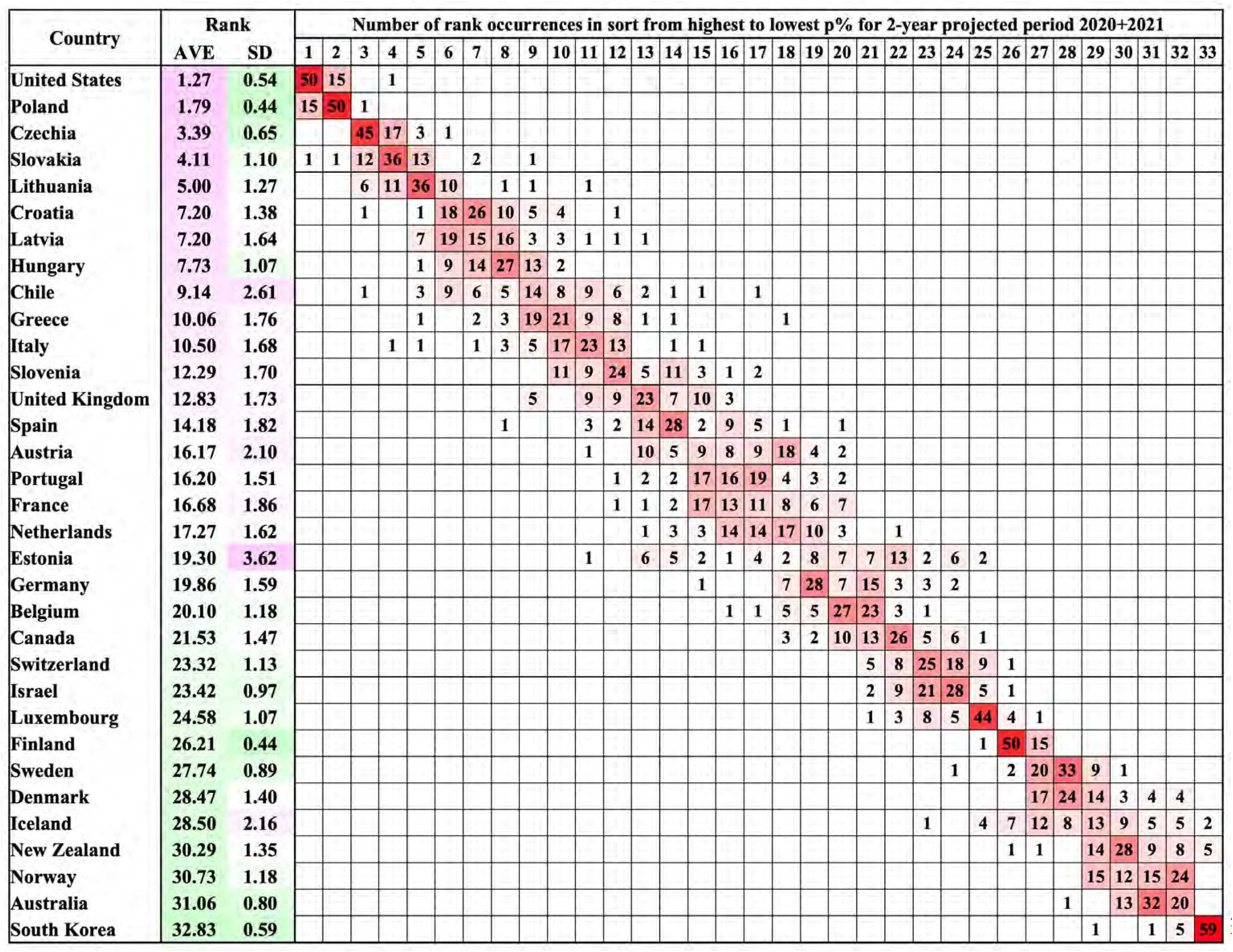
Distribution of the country rank of the excess death estimates (from highest to lowest) in the pandemic 2-year projected period 2020+2021 expressed as a percentage of the expected deaths for the 33 countries as calculated for each of the 66 different reference baseline year sets. The countries are ordered by decreasing average rank (column 3); the standard deviation of the rank is given in column 4. For 23 countries, the most common occurrence is on the diagonal. For the 6 countries between Austria and Germany, the average rank is between 16.2 and 19.9 and the rank order is ambiguous. For 21 countries the most common rank occurrence is on the diagonal.

### Diversity in time patterns across 33 countries

Figure 2 maps the emerging time patterns for mortality in each of the analyzed countries for the average of the 66 analyses using different reference baseline periods and the range of maximum and minimum estimates. Although the range of estimates of relative mortality for each given year is large, the rank of different years for a particular country is generally the same for the 66 different sets of reference years (Figure S3). Time patterns across different countries show large variability as well. Differences exist both in the presence and magnitude/steepness of time trends; and on the presence or not of peaks of mortality impact during the COVID-19 pandemic (2020, 2021, both, or neither). All countries had some decline in mortality over the period 2009-2019, but for the USA in particular the change was minimal (change from average of 1.27% in 2009-2010 to -1.31% in 2018-2019 for an overall decline of only 2.58%, using data in Table S1B). The other 4 countries with the smallest changes for the averages between 2009-2010 and 2018-2019 were Germany, The Netherlands, the United Kingdom and Canada , (changes of -6.65%, -8.34%, -8.49% and -8.55%, respectively). Conversely, the 5 countries with the largest declines for the averages between 2009-2010 and 2018-2019 were South Korea, Estonia, Denmark, Slovakia and Norway (changes of -23.3%, -19.7%, -17.1%, -16.0% and - 14.1%, respectively). For the pandemic period 2020-2021, the USA had the steepest increase (change in average 18.00% between 2018-2019 and 2020-2021). Steep increases were seen also in Eastern European and Balkan countries (changes in average from 10.18% to 17.46% between 2018-2019 and 2020-2021 for Slovenia, Hungary, Latvia, Croatia, Lithuania, Czechia, Slovakia and Poland). Most western European countries had more modest disruptions of the declining trend (changes for the averages from 1.86% to 9.98% between 2018-2019 and 2020-2021 for Luxembourg, Germany, Switzerland, France, The Netherlands, Belgium, Portugal, Austria, the United Kingdom, Spain and Italy). Some Scandinavian countries, Australia, New Zealand, and South Korea continued to have declining mortality trends during the pandemic (changes for the averages from -4.97% to -2.44% between 2018-2019 and 2020-2021 for New Zealand, South Korea, Iceland, Norway, Denmark and Australia). Figures S4A, S4B, and S4C map the time patterns shown in Figure S2 for periods of 1, 3 and 4 years, respectively and show how longer projected periods reduce fluctuations.

**Figure 2:**
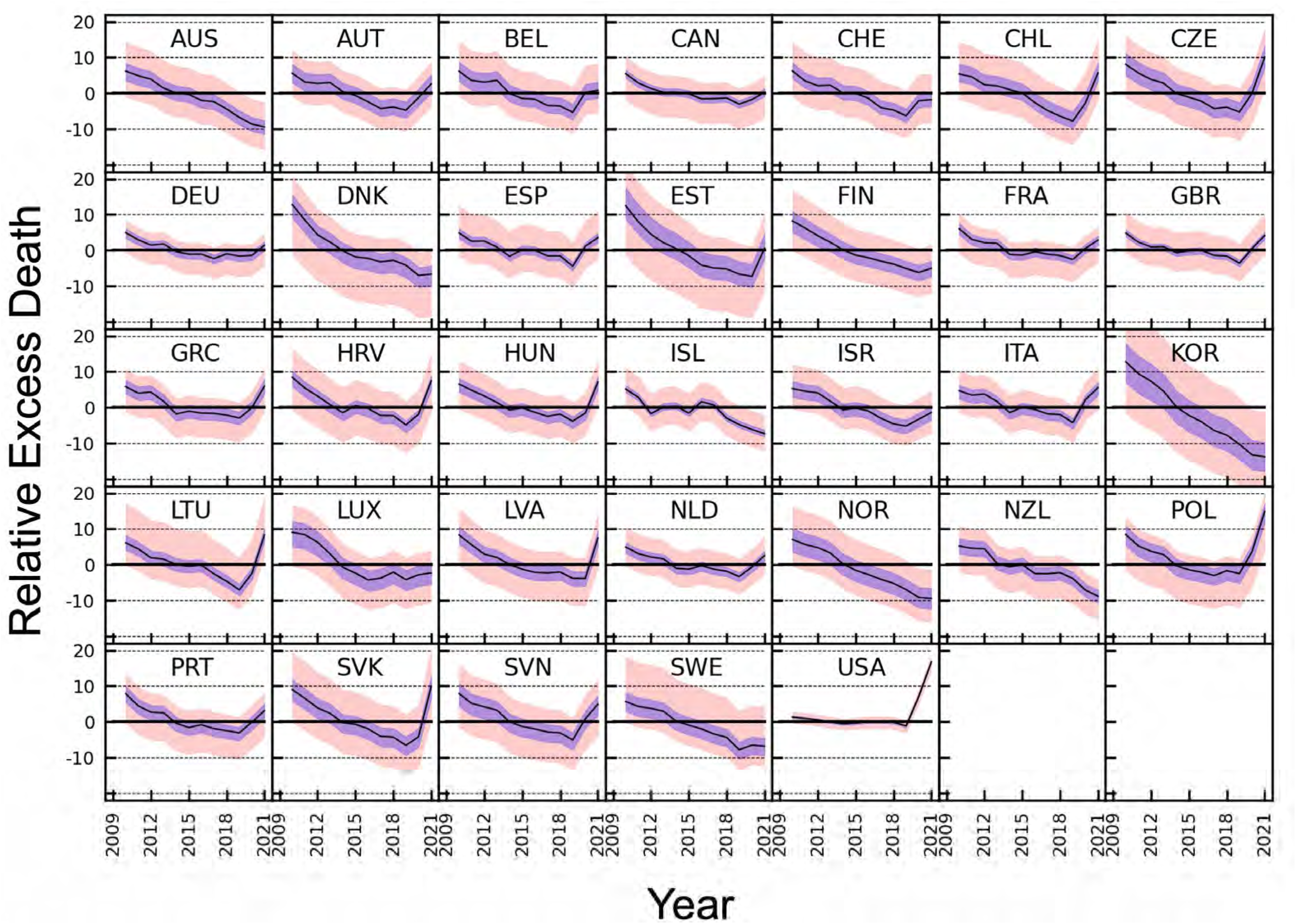
Variation with year from 2009 to 2021 of the excess death estimates expressed as a percentage of the expected deaths. The expected deaths are estimated from the average mortality values of each of the 66 different reference year-sets, which are all combinations of one or more consecutive years from 2009 to 2019. The y-axis of every panel extends from -22% to 22%. The plots for different reference year sets are almost identical but shifted along the y-axis by different amounts. The two year predicted period, which is particularly significant as the complete pandemic years are 2020+2021, is shown here; other projected periods with 1, 3 and 4 years are shown in Figures S4 A, B &C. The salmon shading marks the range of all 66 reference periods, the purple shading marks the range between the first and third quartile and the black shows the median for the reference periods. The 3-letter country abbreviations are: AUS: Australia, AUT: Austria, BEL: Belgium, CAN: Canada, CHE: Switzerland, CHL: Chile, CZE: Czechia, DEU: Germany, DNK: Denmark, ESP: Spain, EST: Estonia, FIN: Finland, FRA: France, GBR: United Kingdom, GRC: Greece, HRV: Croatia, HUN: Hungary, ISL: Iceland, ISR: Israel, ITA: Italy, KOR: South Korea, LTU: Lithuania, LUX: Luxembourg, LVA: Latvia, NLD: Netherlands, NOR: Norway, NZL: New Zealand, POL: Poland, PRT: Portugal, SVK: Slovakia, SVN: Slovenia, SWE: Sweden, and USA: United States.

Table S2 and Tables S1A,B,C,D present data on the worst years. Table S1A shows that the worst single year with the highest mortality was 2021 for 10 countries (Slovakia, Poland, United States, Latvia, Lithuania, Hungary, Croatia, Czechia, Chile and Greece), 2020 was the highest for 4 countries (United Kingdom, Italy, Spain and Belgium), 2010 was worst for Luxembourg and 2009 was worst for all other 18 countries. When considering 2-year periods, in 25 of the 33 countries, 2009+2010 were the worst pair of years (Table S2). In 9 of the 33 countries (Chile, Czechia, Greece, Hungary, Italy, Lithuania, Poland, Slovakia and United States) the pandemic years 2020+2021 were the worst, and in all of them the years 2009+2010 were the second worst. (Table S1A & Table 3). In 16 countries, the pandemic years were not among the three worst years, which were always years between 2009 and 2016. When considering 3 or 4 year periods, in 31 of the 33 countries 2009-2011 and 2009-2012, were the worst, respectively. Only in Poland or the United States were period 2019-2021 and 2018-2021, which include the pandemic years, the worst, respectively, (data from Tables S1C & S1D)

**Table 2:**
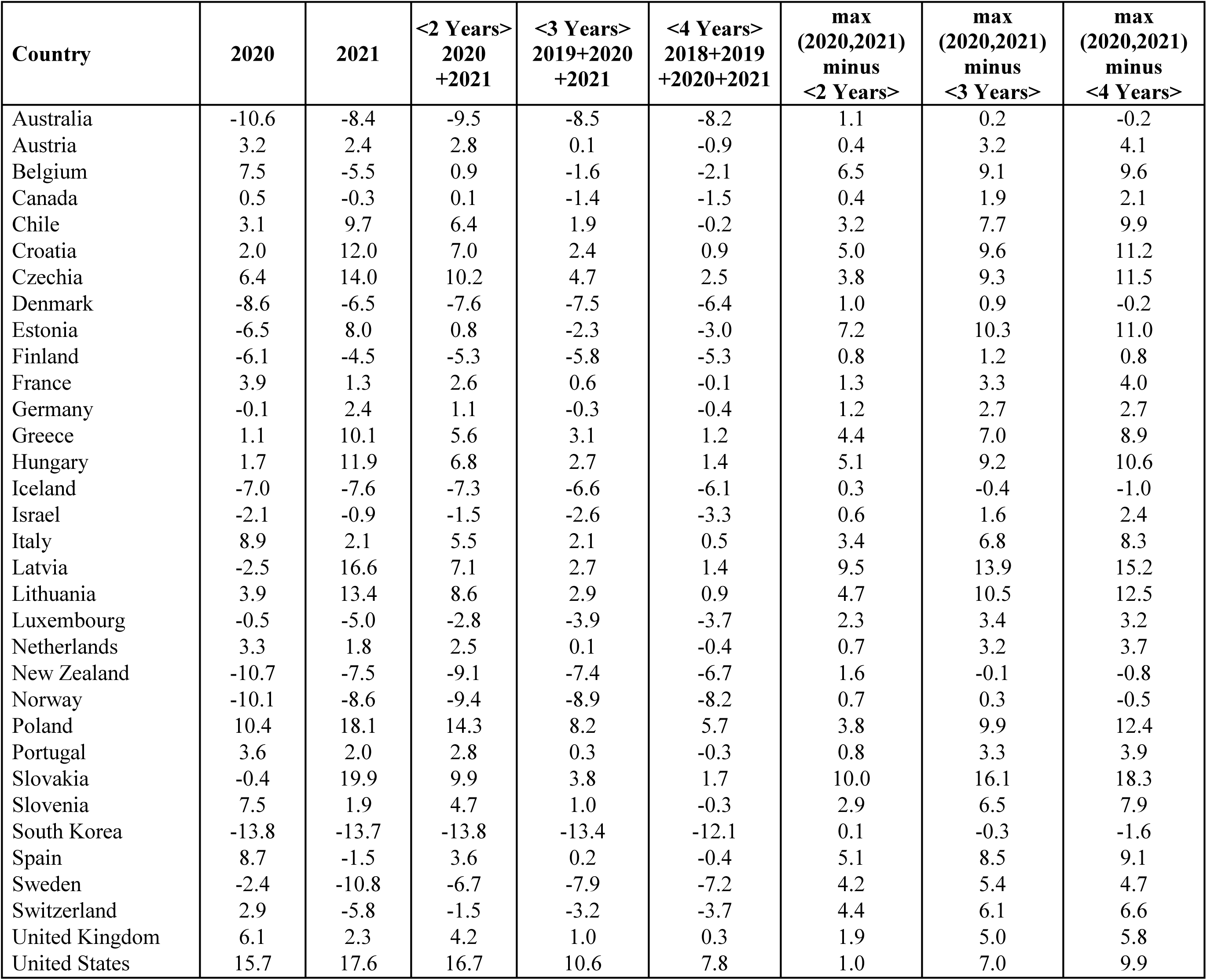
Effect of changing the projected period of interest from 1 to 4 years for the most recent years: 2021 alone, 2020 alone, <2 Years> = 2020+2021, <3 Years> = 2019+2020+2021, and <4 Years> = 2018+2019+2020+2021).

| **Country** | **2020** | **2021** | **<2 Years>**<br>**2020**<br>**+2021** | **<3 Years>**<br>**2019+2020**<br>**+2021** | **<4 Years>**<br>**2018+2019**<br>**+2020+2021** | **max**<br>**(2020,2021)**<br>**minus**<br>**<2 Years>** | **max**<br>**(2020,2021)**<br>**minus**<br>**<3 Years>** | **max**<br>**(2020,2021)**<br>**minus**<br>**<4 Years>** |
| --- | --- | --- | --- | --- | --- | --- | --- | --- |
| Australia | -10.6 | -8.4 | -9.5 | -8.5 | -8.2 | 1.1 | 0.2 | -0.2 |
| Austria | 3.2 | 2.4 | 2.8 | 0.1 | -0.9 | 0.4 | 3.2 | 4.1 |
| Belgium | 7.5 | -5.5 | 0.9 | -1.6 | -2.1 | 6.5 | 9.1 | 9.6 |
| Canada | 0.5 | -0.3 | 0.1 | -1.4 | -1.5 | 0.4 | 1.9 | 2.1 |
| Chile | 3.1 | 9.7 | 6.4 | 1.9 | -0.2 | 3.2 | 7.7 | 9.9 |
| Croatia | 2.0 | 12.0 | 7.0 | 2.4 | 0.9 | 5.0 | 9.6 | 11.2 |
| Czechia | 6.4 | 14.0 | 10.2 | 4.7 | 2.5 | 3.8 | 9.3 | 11.5 |
| Denmark | -8.6 | -6.5 | -7.6 | -7.5 | -6.4 | 1.0 | 0.9 | -0.2 |
| Estonia | -6.5 | 8.0 | 0.8 | -2.3 | -3.0 | 7.2 | 10.3 | 11.0 |
| Finland | -6.1 | -4.5 | -5.3 | -5.8 | -5.3 | 0.8 | 1.2 | 0.8 |
| France | 3.9 | 1.3 | 2.6 | 0.6 | -0.1 | 1.3 | 3.3 | 4.0 |
| Germany | -0.1 | 2.4 | 1.1 | -0.3 | -0.4 | 1.2 | 2.7 | 2.7 |
| Greece | 1.1 | 10.1 | 5.6 | 3.1 | 1.2 | 4.4 | 7.0 | 8.9 |
| Hungary | 1.7 | 11.9 | 6.8 | 2.7 | 1.4 | 5.1 | 9.2 | 10.6 |
| Iceland | -7.0 | -7.6 | -7.3 | -6.6 | -6.1 | 0.3 | -0.4 | -1.0 |
| Israel | -2.1 | -0.9 | -1.5 | -2.6 | -3.3 | 0.6 | 1.6 | 2.4 |
| Italy | 8.9 | 2.1 | 5.5 | 2.1 | 0.5 | 3.4 | 6.8 | 8.3 |
| Latvia | -2.5 | 16.6 | 7.1 | 2.7 | 1.4 | 9.5 | 13.9 | 15.2 |
| Lithuania | 3.9 | 13.4 | 8.6 | 2.9 | 0.9 | 4.7 | 10.5 | 12.5 |
| Luxembourg | -0.5 | -5.0 | -2.8 | -3.9 | -3.7 | 2.3 | 3.4 | 3.2 |
| Netherlands | 3.3 | 1.8 | 2.5 | 0.1 | -0.4 | 0.7 | 3.2 | 3.7 |
| New Zealand | -10.7 | -7.5 | -9.1 | -7.4 | -6.7 | 1.6 | -0.1 | -0.8 |
| Norway | -10.1 | -8.6 | -9.4 | -8.9 | -8.2 | 0.7 | 0.3 | -0.5 |
| Poland | 10.4 | 18.1 | 14.3 | 8.2 | 5.7 | 3.8 | 9.9 | 12.4 |
| Portugal | 3.6 | 2.0 | 2.8 | 0.3 | -0.3 | 0.8 | 3.3 | 3.9 |
| Slovakia | -0.4 | 19.9 | 9.9 | 3.8 | 1.7 | 10.0 | 16.1 | 18.3 |
| Slovenia | 7.5 | 1.9 | 4.7 | 1.0 | -0.3 | 2.9 | 6.5 | 7.9 |
| South Korea | -13.8 | -13.7 | -13.8 | -13.4 | -12.1 | 0.1 | -0.3 | -1.6 |
| Spain | 8.7 | -1.5 | 3.6 | 0.2 | -0.4 | 5.1 | 8.5 | 9.1 |
| Sweden | -2.4 | -10.8 | -6.7 | -7.9 | -7.2 | 4.2 | 5.4 | 4.7 |
| Switzerland | 2.9 | -5.8 | -1.5 | -3.2 | -3.7 | 4.4 | 6.1 | 6.6 |
| United Kingdom | 6.1 | 2.3 | 4.2 | 1.0 | 0.3 | 1.9 | 5.0 | 5.8 |
| United States | 15.7 | 17.6 | 16.7 | 10.6 | 7.8 | 1.0 | 7.0 | 9.9 |

### Excess death estimates in recent years using different projected period time windows

Table 2 shows the effect of changing the width of the projected period of interest from 1 to 4 years for the most recent years (2021 alone, 2020 alone, 2020-2021, 2019-2021, 2018-2021). As shown, there is substantial attenuation of the relative excess mortality between the single worse pandemic year and increasingly wider periods of interest. The attenuation was most prominent when averaging over a 4-year period for Slovakia, Latvia, Lithuania, Poland, Estonia and Croatia, with relative drops of 18.3, 15.2, 12.5, 12.4, 11.0 and 11.2 percentage points, respectively. The attenuation was least prominent for Australia, Norway, Denmark, Iceland, New Zealand and South Korea, with relative drops of -0.2, -0.5, -0.20, -1.0, -0.8 and -1.6, percentage points, respectively. The USA maintained the most prominent peak even with a 4-year window.

With increasing projected periods, both the mean and standard deviation of the relative excess mortality declined substantially. For 2021, 2020-2021, 2019-2021, 2018-2021, the mean was 2.6%, 1.5%, -1.0%, and -1.7%, respectively. The standard deviation was 9.3%, 7.2%, 5.2%, and 4.1%, respectively.

## DISCUSSION

Our application of a multiverse approach to excess death data shows that consideration of different periods for reference baseline resulted in major variability in the absolute magnitude of the exact excess death estimates, but it did not affect substantially the relative ranking of different countries compared to others for specific years. Moreover, there have been distinct time patterns across different countries during 2009-2021. Countries differed markedly on whether they had a substantial decrease over time or not during 2009-2021, on whether they had a peak during the 2020-2021 pandemic years, and, if so, how high, and in the relative contribution of 2020 and of 2021 to this peak. With longer time windows for the projected period of interest (1 to 4 years), the range of excess deaths across different countries in the pandemic years and the 2 years preceding the pandemic shrank substantially and excess death estimates became less variable across countries. This suggests that it would be inappropriate to dwell too much on small or modest differences between countries, as these are highly model-dependent. However, peaks did not disappear and for the USA in particular, excess deaths remained prominent even with long projected periods of interest.

In the multiverse literature from other fields, some analytical choices may be considered more meaningful or relevant than others. When researchers are asked to select independently what analysis mode they feel is most sensible, not all analytical choices are selected (19,20) and some types of choices may seem to make more sense. This may apply also for excess death calculations. E.g. it may seem not so appropriate to use a reference window of 2009 alone for projecting mortality in 2021. The baselines created by each of the 66 different windows may have less or more relevance to the current situation. In principle, baselines using more recent years may be more informative for the current time. Common choices include using the last 3 years or the last 5 years. However, examining all possible baselines allows to reveal in a systematic manner any long-term patterns in mortality.

The obvious heterogeneity of time patterns across different countries suggests that selection of specific time trends in modeling excess deaths may be a situation where one size does not fit all. Selection of specific anticipated time trend patterns may markedly affect the results in ways that are not verifiable for their appropriateness. E.g. selecting a model that anticipates a marked decrease in mortality over time makes it difficult for a country not to have excess deaths even if it does very well in a given year – but still falls short of an anticipated stellar improvement over time. It should be acknowledged that age-adjusted mortality rates usually have decreased over time in most countries in the last several decades. Standard methods for forecasting future mortality rates and life expectancy such as the Lee-Carter forecasts (22,23) and other methods that use time series approaches end up using some linear trends in the modeling. However, it has been observed (24) that changes in mortality rates may differ markedly in different years even in the same country/location and they may also differ across different age and gender groups in the same country and same year. In the presence of major perturbation events such as pandemics or wars and natural disasters, such modeling will unavoidably fail. More importantly, there is no guarantee that mortality rates should continue declining, let alone markedly decline, over time with medical and other progress, even in the absence of major negative perturbation events (6). For advanced economies with aging populations, accumulating frailty and disease burden and restrictions or ceilings to progress and available resources, the typical trends for decreased mortality that were documented in the previous decades may not be sustainable for the future. Furthermore, countries that have already reached very high life expectancies may have less room for improvement than others that are lagging behind. The multiverse approach, when applied to multiple countries, allows a comparative assessment of the trajectory of different countries. This may be preferable and it may offer some genuine insights about which countries do well (short-term and long-term) and which do poorly – in comparison.

In this regard, some stark differences stand out for both long-term trends and for the pandemic years. The USA consistently performed very poorly with both stagnation in mortality during the pre-pandemic years and a sharp increase during the pandemic. Eastern European and Balkan countries showed sharp decreases during the pre-pandemic years and a sharp increase during the pandemic. Most western European countries had sharp decreasing trends with modest disruption during the pandemic. All Scandinavian countries, Australia, New Zealand, and South Korea have had largely unperturbed declining mortality patterns. The markedly different patterns may reflect a combination of social, health care, and pandemic factors. The USA has an ailing health system with approximately 30 million uninsured people (25), large inequalities (26), many people with poor access to care (27), and major ongoing non-infectious epidemics, including obesity (28), opioid abuse and overdose (29), and violent deaths (30). More detailed data are needed to understand which of the policies and actions during the pandemic or the pre-existing problems were more important for shaping the poor performance in 2020 and beyond.

Eastern European and Balkan countries have limited resources for their healthcare systems and lower social welfare than other European countries (31) and some countries like Greece have long suffered from austerity (32). The best performers are excelling in social welfare and health system functionality and resources, even if there are differences across countries. Exceptions may occur within circumscribed populations and adverse settings even in countries with overall excellent trajectories. For example, the dysfunctional consequences of privatization in nursing homes in countries like Sweden or Canada (33,34) translated to peaks of excess deaths during circumscribed periods in the long-term care settings (35).

Consideration of longer projected period time windows diminished substantially the range of excess deaths in some countries, but not in others. Overall, when longer periods are considered, differences between most countries become less pronounced. However, larger windows had minimal effect in the USA, and this may reflect that the problems that lead to unfavorable mortality patterns in the USA reflect chronic dysfunctions that might have been accentuated by the pandemic but pre-existed and which affect also people with long life expectancy. Poverty, marginalization, homelessness, inequalities, drug overdoses, and violence affect indeed young and middle-aged populations. We have shown previously that the USA has had 40% of excess deaths contributed by the <65 age stratum, a higher percentage than all other highly developed countries (6). Conversely, in many other countries, large time windows for the projected period shrank substantially the excess death fluctuations. This suggests that in these countries excess deaths temporarily affect mostly people with relatively limited life expectancy (21).

Europe, while not a country, has historically aggregated excess death data in the EuroMOMO data base (https://www.euromomo.eu/) to include data from 21 countries in Europe plus Israel (36). If one were to aggregate data for the 19 of these 21 countries for which we have data (excluding Cyprus & Ireland), the fictional country composite that includes Austria, Belgium, Denmark, Estonia, Finland, France, Germany, Greece, Hungary, Israel, Italy, Luxembourg, Netherlands, Norway, Portugal, Slovenia, Sweden, Switzerland, and United Kingdom has a similar population to the USA (410 million versus 330 million) and the relative excess death of this European composite is only 2.46% for the pandemic years (2020+2021), which is in stark difference to the USA figures. It is also less than two-year totals for 2009+2010 and 2010+2011, with values of 5.57% and 3.30% caused by elevated Influenza pandemics (37). One may examine also the excess deaths according to the EuroMOMO model, but the model has been criticized for low baseline values which may lead to overestimation of excess mortality in some countries (38).

Some limitations should be acknowledged. First, there are some additional sources of analytical flexibility that can be considered in excess death calculations. These include the choice of age bins for age adjustment, and the use of additional adjustments for modeling the population profile over time. For example, socioeconomic profile variables would be very useful to incorporate (39), but these are not routinely available and standardized across many countries. Such additional adjustments would add additional variation with more multiverse options, but probably would not invalidate the major patterns that we observed. Second, we only modeled data from 33 countries that are the ones with the most reliable data. Extrapolations to other countries would be precarious, given the unreliability of the mortality information. Time patterns observed in the 33 countries may not necessarily apply to the remaining countries around the globe and local circumstances may make a difference. Third, we considered yearly interval increments so as to capture all 4 seasons in the unit of time, but in theory, the multiverse process can be applied for smaller units of time as well. Fourth, data on population and population structure in each country on a yearly basis are typically inferred from census data collected on more sparse timing, therefore they carry some uncertainty. Fifth, the pandemic impact and its consequences as well as the consequences of aggressive measures that were taken has continued more prominently in 2022 in some countries than others (40). It would be interesting to see whether differences across countries get further attenuated and/or some countries continue to stand out prominently when longer pandemic and post-pandemic periods (e.g. 2020-2022 and 2020-2023) are considered. Preliminarily results based on the first 8 months of 2022, it seems that several countries with death deficit in 2020-2021 (e.g., Australia, New Zealand and South Korea), had considerable excess deaths in 2022, while some others continued to have limited deaths (e.g. Sweden) and some hard-hit countries like USA and Greece continued to do very poorly (6,40). Sixth, we did not consider in the multiverse analyses any superimposed modeling of time trends, specifically because we wanted to allow the data to show whatever trends of patterns existed. If one were to add in the modeling also all the possible functions that might be used to capture time trends (e.g. linear, higher power, splines, and so forth), the analytical options would multiply far more. This explains why mortality forecasting is so difficult and uncertain, and why there is no consensus on the best method on how to do it (41).

Mortality forecasting becomes even more difficult and uncertain when perturbation events such as pandemics occur. The multiverse approach helps understand why obtaining accurate absolute estimates of excess deaths is precarious. However, our approach using down weighted older reference years may be considered, if absolute estimates are desirable (as opposed to relative performance across years and compared with other countries). One may also down weight previous reference years based on other features, e.g. severe flu seasons or major heat waves.

In conclusion, a multiverse approach to excess death calculations may offer bird’s eye views on mortality patterns in comparative assessments of a large number of countries. These patterns may be more reliably informative than efforts to obtain isolated single-country estimates of excess deaths, which are subject to substantial uncertainty even in countries with the best-collected data. It may be best to avoid pre-specifying time patterns and to allow the data to show what time patterns may be emerging. Finally, observed time patterns may not necessarily continue into the future and multiverse analyses can be updated accordingly for additional years moving forward.

## MATERIALS AND METHODS

### Data

All data comes from the Human Mortality Database (HMD) (42–44). The data for the most recent years comes from the Short-Term Mortality Fluctuation file stmf.csv downloaded from https://www.mortality.org/File/GetDocument/Public/STMF/Outputs/stmf.csv (last updated 6 February 2023). The data for earlier years extending back to 2009 was downloaded as the HMD archive file (see Supplementary Links to Data). We considered data from 2009-2021 so as to analyze 13 years including also the years of the 2009-2020 pandemic. We focused on the 33 high-income countries with highly reliable death registration systems, excluding Bulgaria as done in previous work (6). The most recent data in the file stmf.csv is per week and uses five standard age-bands: 0-14, 15-64, 65-74, 75-84 and Over 85; we sum the data over the weeks assigned to each year as done before (6). The older data in the HMD archive (downloaded on May 25, 2022) uses 1-year age bands for annual all-cause deaths and annual populations are available for all 33 countries. We sum these 1-year bands to give the same five standard age bands used in stmf.csv.

### Excess death calculations

In order to be able to compare different countries and different time periods we focus on relative excess deaths expressed as the number of excess deaths divided by the number of expected deaths. Specifically, the relative excess death p% is the actual all-cause death count, D, minus the estimated death count, E, expressed as a percentage of the estimated death count or *p%=(D-E)/E*.

### Systematic Variation of Assumptions for Multiverse Analyses

We consider all possible reference baseline periods and projected periods in consecutive year time windows during a lengthy period of interest. Instead of prespecifying time patterns, this multiverse approach allows the data to demonstrate what might be the time patterns and how sensitive the results are to different analytical choices. No linear or spline or other trends are imposed on the data; instead, such trends are allowed to be revealed by the patterns shown by using all possible averages of reference years as the baseline.

We consider all possible reference baseline spans of consecutive years in the period 2009-2019. This gives 11+10+..+1=66 different spans of length 1 to 11 years for the 66 reference baselines. For each reference period, we average the mortality of each of the five age bands. These averaged mortalities are then used to get the expected deaths in any year by multiplying the mortality of a particular age band by the population of that age band and then summing the estimated values over the five age bands to give the total estimated death count.

Projected time periods are also considered in all possible options of length 1 to 4 years, again considering consecutive calendar years. The mortality in the pandemic years 2020 and 2021 is never considered when calculating excess death. Similarly, when calculating excess deaths for projected periods 2018+2019+2020+2021 (or 2019+2020+2021), the years 2018-2019 (or 2019) are not considered as baselines. For analyses with different assumptions, we present the maximum, minimum, median and IQR or mean and standard deviation, as appropriate).

### Analyses with down weighting for older years

Estimates of excess deaths during the pandemic years that are averaged against all possible reference periods may be misleading in absolute magnitude, since very early years such as 2009 may not be as relevant as more recent years like 2019. Therefore, we also rerun the analyses for all 66 possible combinations of reference years with various weights: (a) with weights decreasing linearly by 10% for each year before 2019 (i.e. 100% weight for 2019, 90% weight for 2018, …, 10% weight for 2010, 0% weight for 2009); (b) with weights decreasing by 5% for each year before 2019 (i.e. 100% weight for 2019, 95% weight for 2018, …, 55% for 2010, 50% for 2009); (c) with weights decreasing by half for each year (i.e. 100% weight for 2019, 50% weight for 2018, 25% weight for 2017, 12.5% weight for 2016, 6.25% weight for 2015, 3.125% weight for 2014, 1.563% for 2013, 0.781% for 2012, 0.390% for 2011, 0.195% for 2010, 0.098% for 2009). In all 3 weighting schemes, the weighted average of the 66 options is obtained, weighting each option by the average weight of the reference years that it contains. For example, the 2018-2019 reference years option is weighted by a factor of 1.9/2=0.95, 1.95/2=0.975, and 1.50/2=0.75, for each of the three weighting schemes above, respectively. The 2017-2019 reference years option is weighted by a factor of 2.7/3=0.9, 2.85/3=0.95, and 1.75/3=0.58, respectively.

## Data availability

All data are in the manuscript, tables, and supplementary tables and in the publicly available databases listed in Supplementary Links to Data and deposited online at https://zenodo.org/record/7095753.

## Data Availability

All data produced in the present study are available upon reasonable request to the authors

https://www.mortality.org/

## ACKNOWLEDGMENTS

None

## 13 SUPPLEMENTARY TABLES & FIGURES

**Figure S1:**
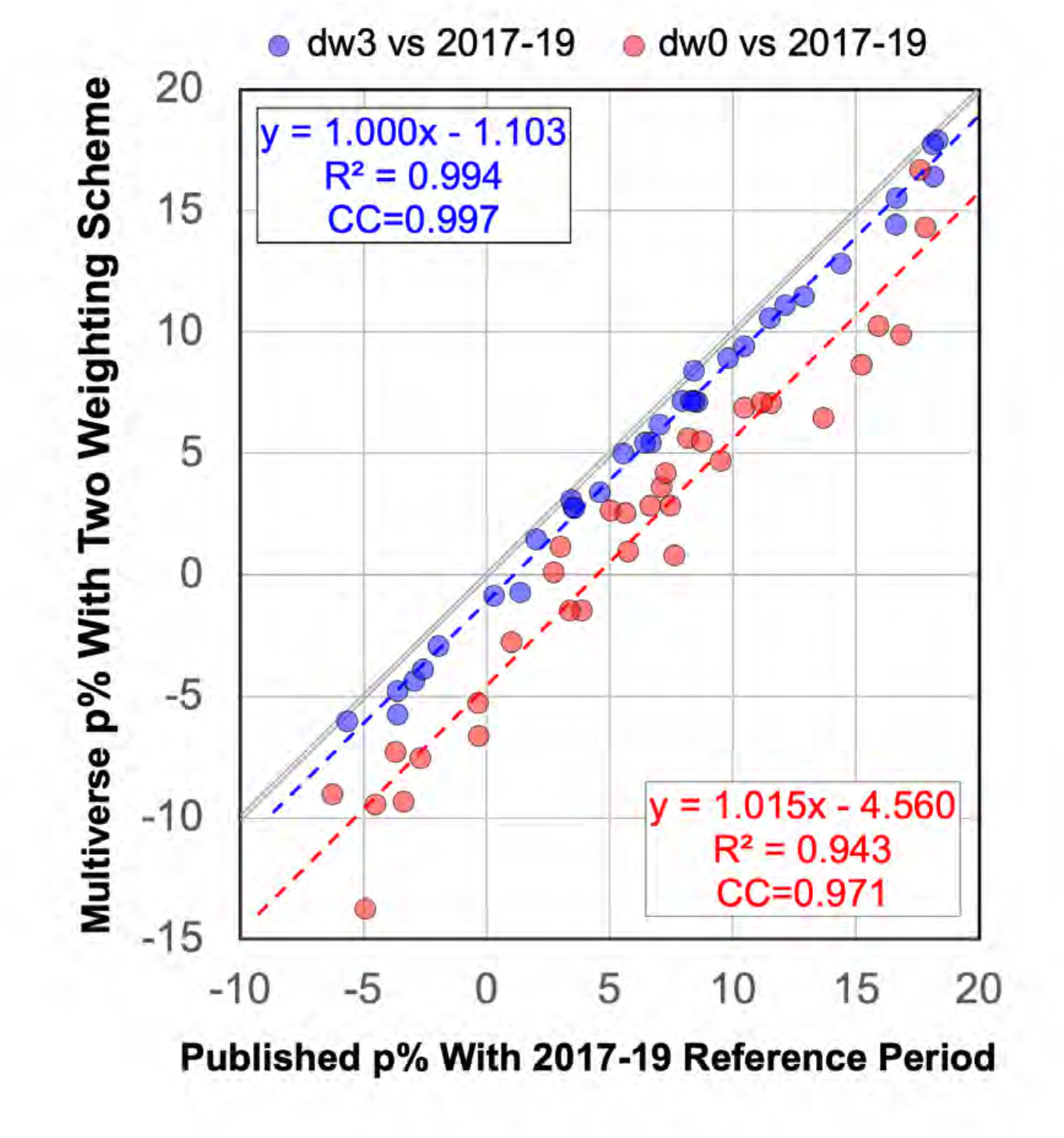
Comparing the multiverse relative excess death, p%, calculated with the 66 reference periods with p% for the single reference period (years 2017 to 2019) published before (6). The multiverse p% values are calculated with dw3 weighting and no weighting (dw0). The fitted linear trends show that dw3 p% values are much closer to 2017-19 p% value with high correlation of 0.997 and average shift of -1.1 percentage points. For the dw0 fit, the correlation is still high at 0.971 but the data is more scattered and the values shift downward by 4.6 percentage points. The gray line is diagonal and shows

**Figure S2A:**
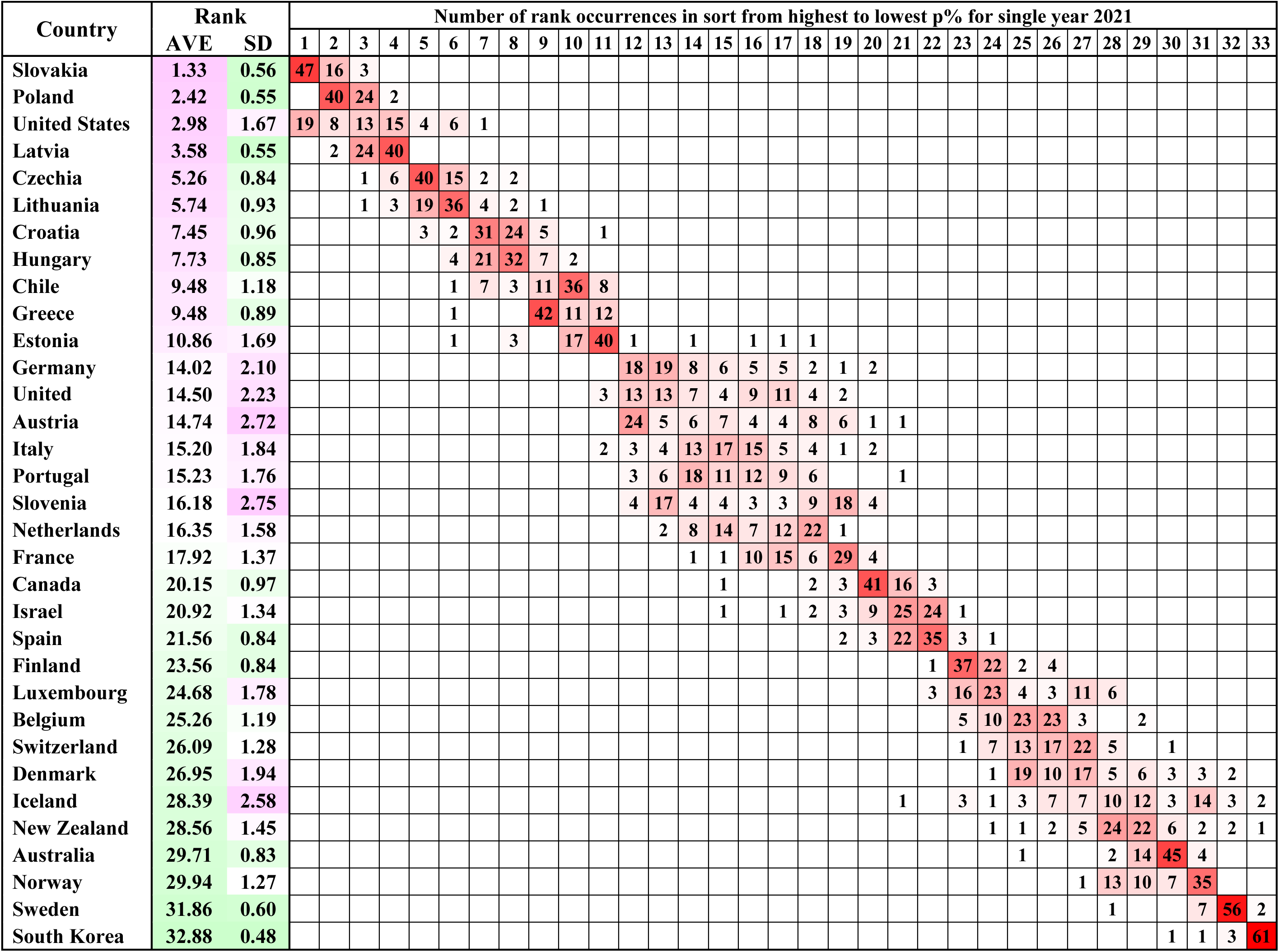
Distribution of the rank of country relative excess death estimates (highest to lowest) in the one-year projected period 2021 for the 33 countries as calculated for each of the 66 different reference baseline year sets. The countries are ordered by decreasing average rank (column 3); the standard deviation of the rank is given in column 4. The country rank is ambiguous for the seven countries between Germany and Netherlands. High- and low-ranking countries are less ambiguous. For 22 countries the diagonal entry occurs most often.

**Figure S2B:**
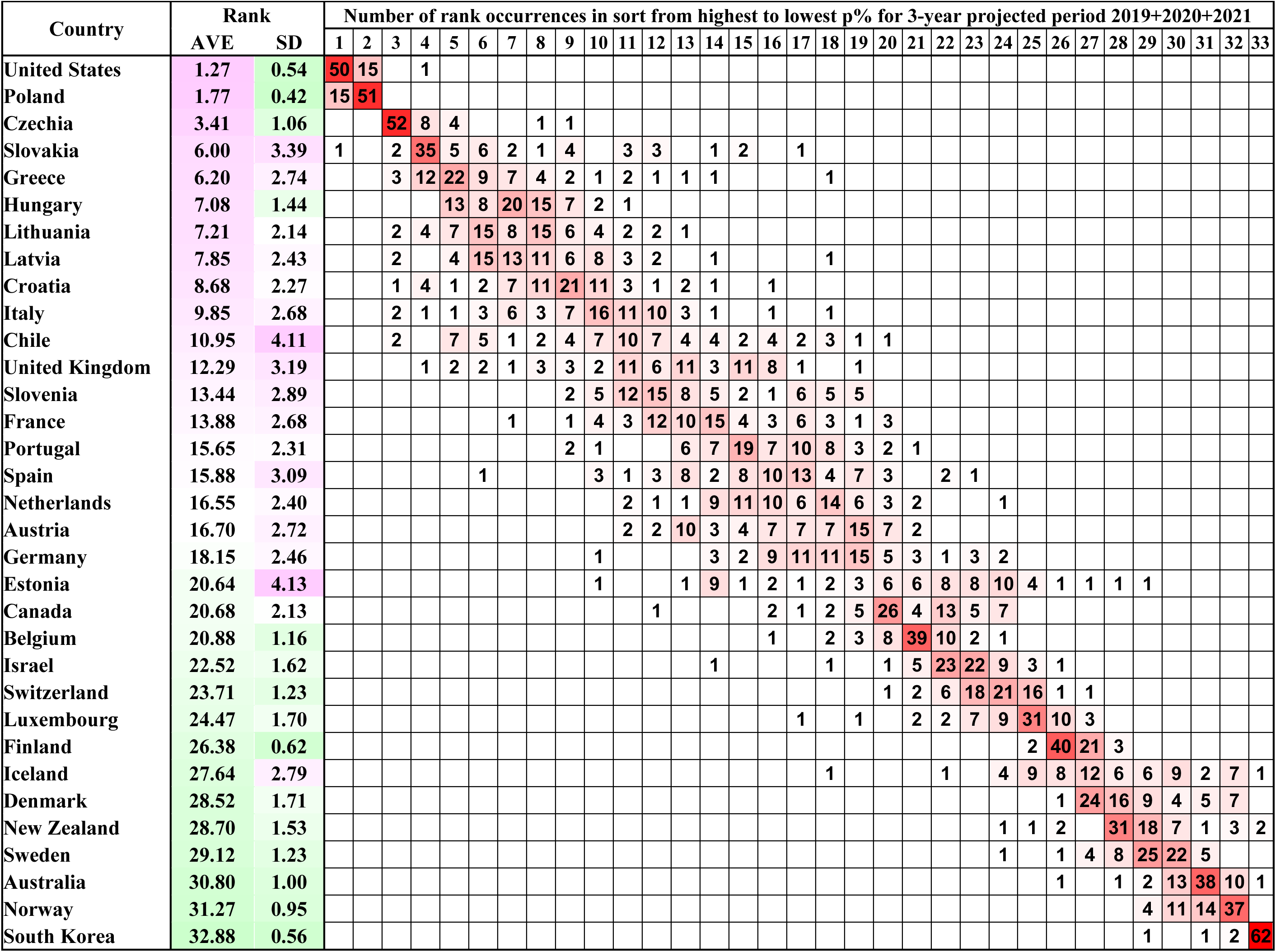
Distribution of the rank of country excess death estimates (from highest to lowest) in the three-year projected period 2019+2020+2021 expressed as a percentage of the expected deaths for the 33 countries as calculated for each of the 66 different reference baseline year sets. The countries are ordered by decreasing average rank (column 3); the standard deviation of the rank is given in column 4. High- and low-ranking countries have consistent ranks but those lying in between are more ambiguous. 18 on diagonal

**Figure S2C:**
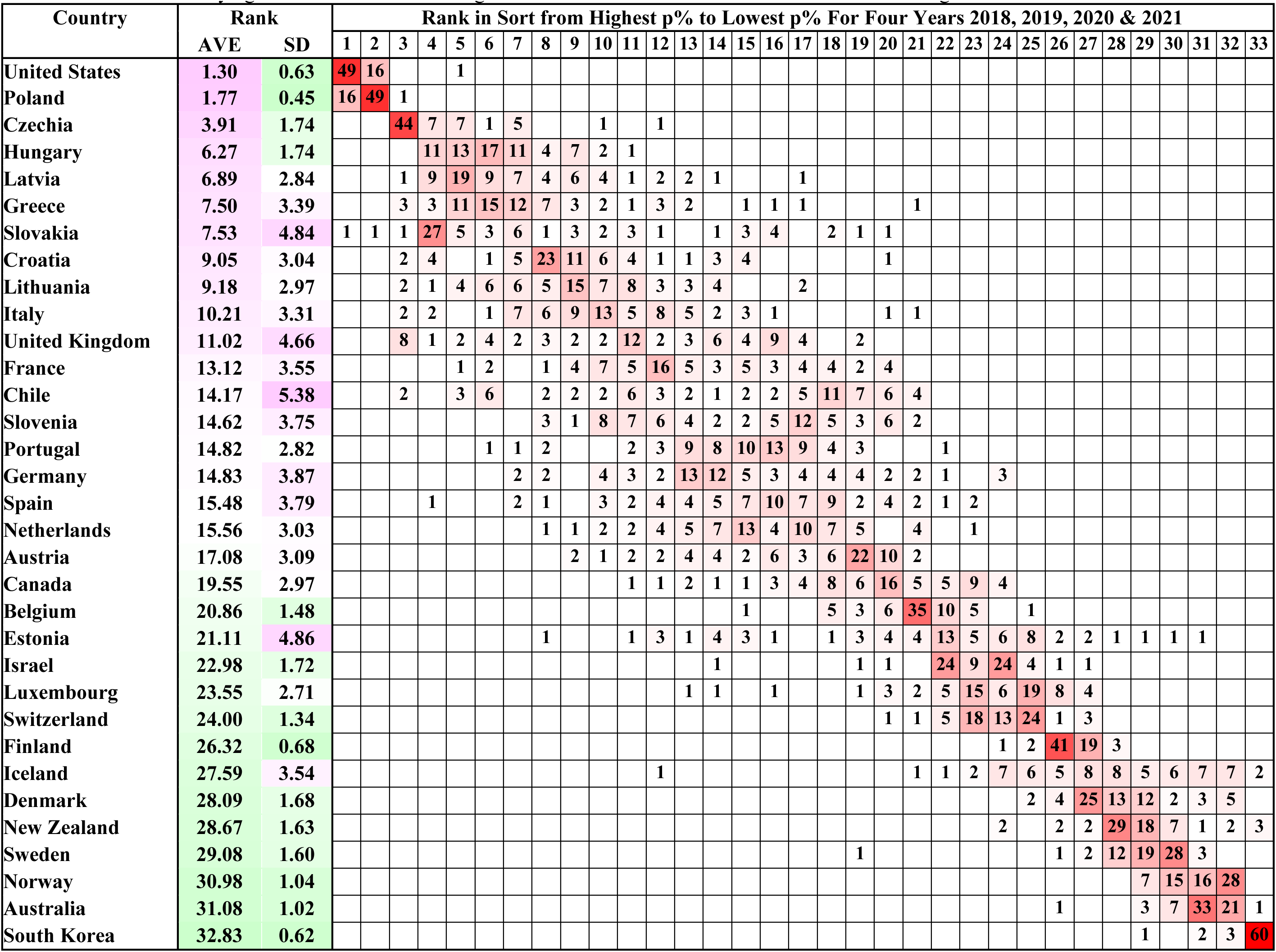
Distribution of the rank of country excess death estimates (from highest to lowest) in the four-year projected period 2018+2019+2020+2021 expressed as a percentage of the expected deaths for the 33 countries as calculated for each of the 66 different reference baseline year sets. The countries are ordered by decreasing average rank (column 3); the standard deviation of the rank is given in column 4. High- and low-ranking countries have consistent ranks but those lying in between are more ambiguous. The most common rank occurrence in on the diagonal 17 times.

**Figure S3:**
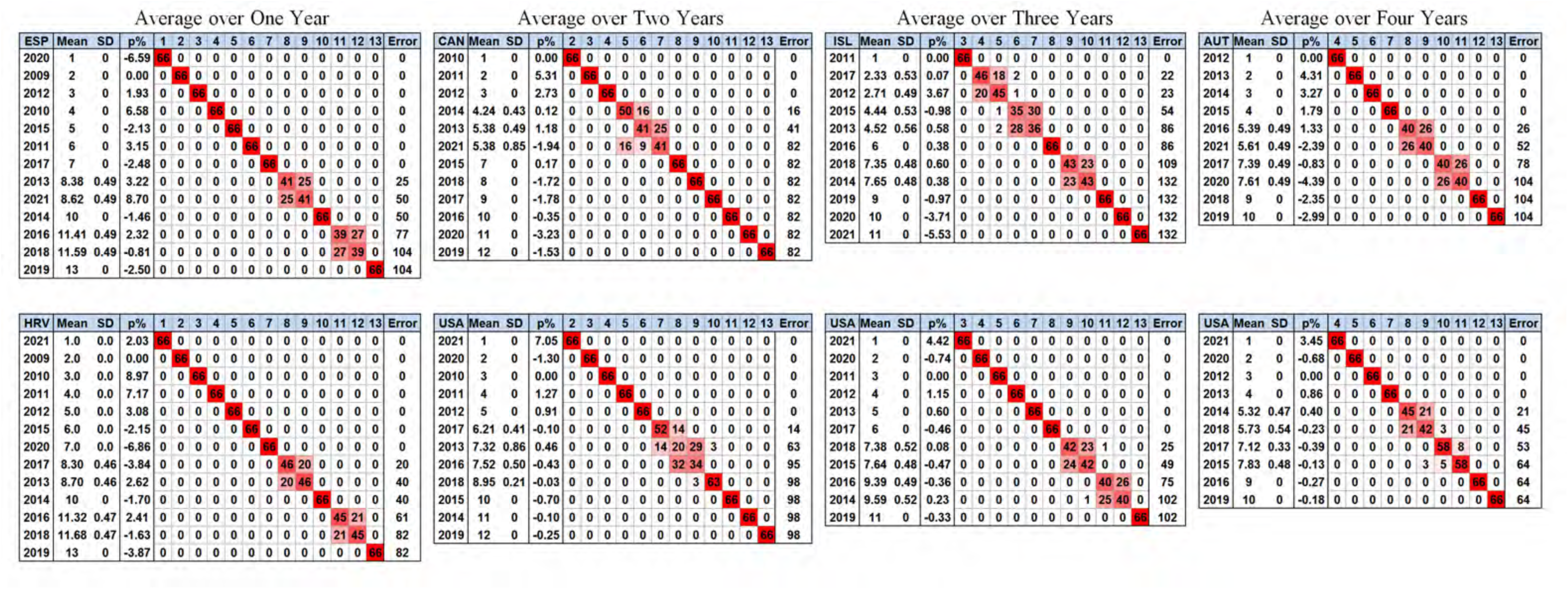
Showing how the ranking of the relative excess death over the years between 2009 and 2021 for the 66 different reference periods is almost independent on the choice of reference set. There are 132 rankings of the 33 countries and 4 averaging periods. Of these 132 rankings, 67 have identical rankings for all reference years. Below we show the two cases for each averaging period with rankings that differ most from the average rank ordering. When not zero, the standard deviation in a ranking is approximately 0.5. Error is defined as the sum of the off-diagonal occurrence multiplied by the distance from the diagonal. Only in one case is the diagonal element smaller than an off-diagonal element: Year 2013 for USA averaged over 2 years and the standard deviation then reaches 0.86. This occurs because the p% values for the USA are flat before 2020 (see Fig. 2) and hard to distinguish.

**Figure S4A:**
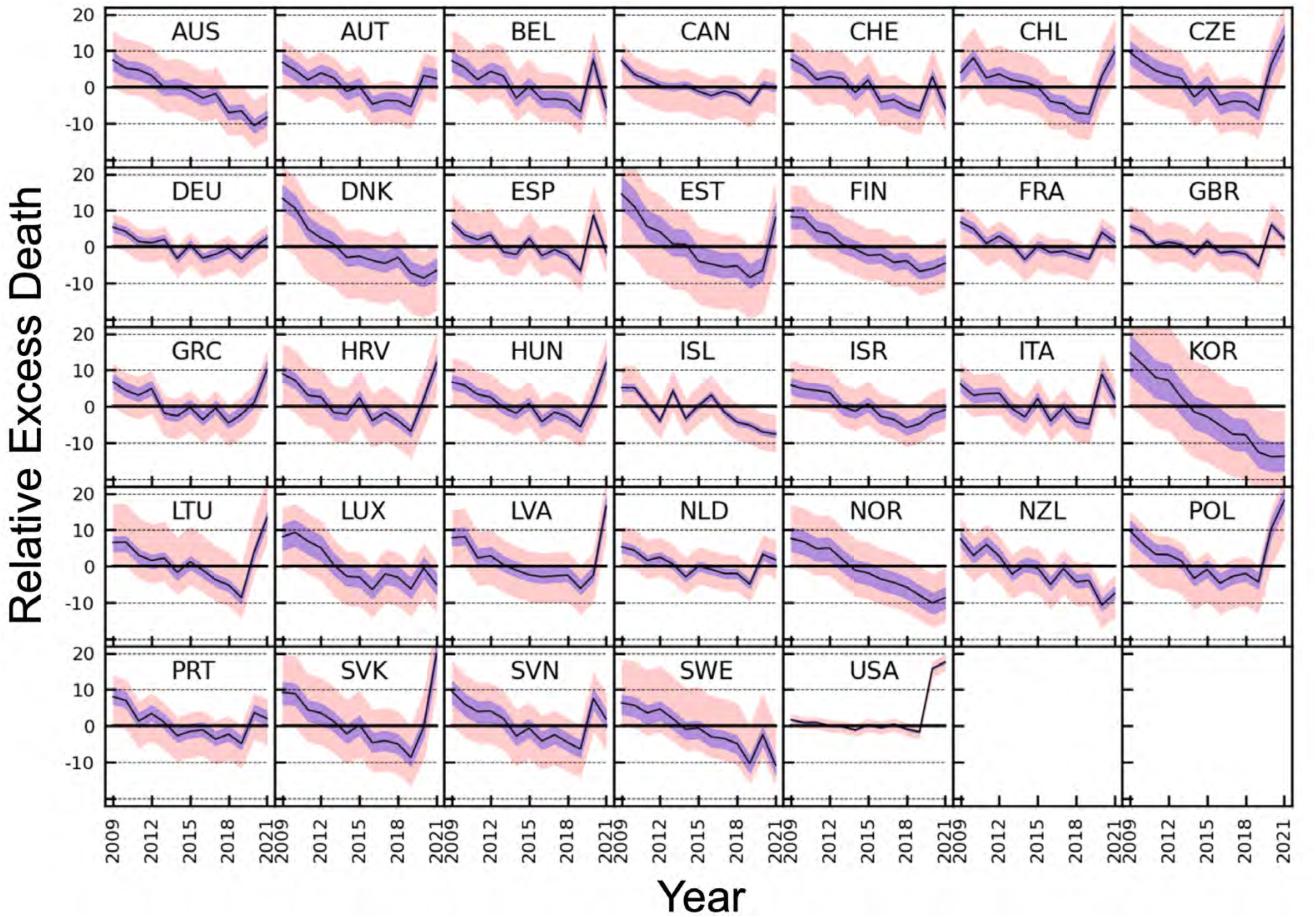
Variation with year from 2009 to 2021 of the relative excess death, p%, calculated for single years. The salmon shading marks the range of all 66 reference periods, the purple shading marks the range between the first and third quartile and the black line shows the median for the reference periods. A year with a low p% value is often followed by a year with a high value. These fluctuations are averaged out with longer projected periods.

**Figure S4B:**
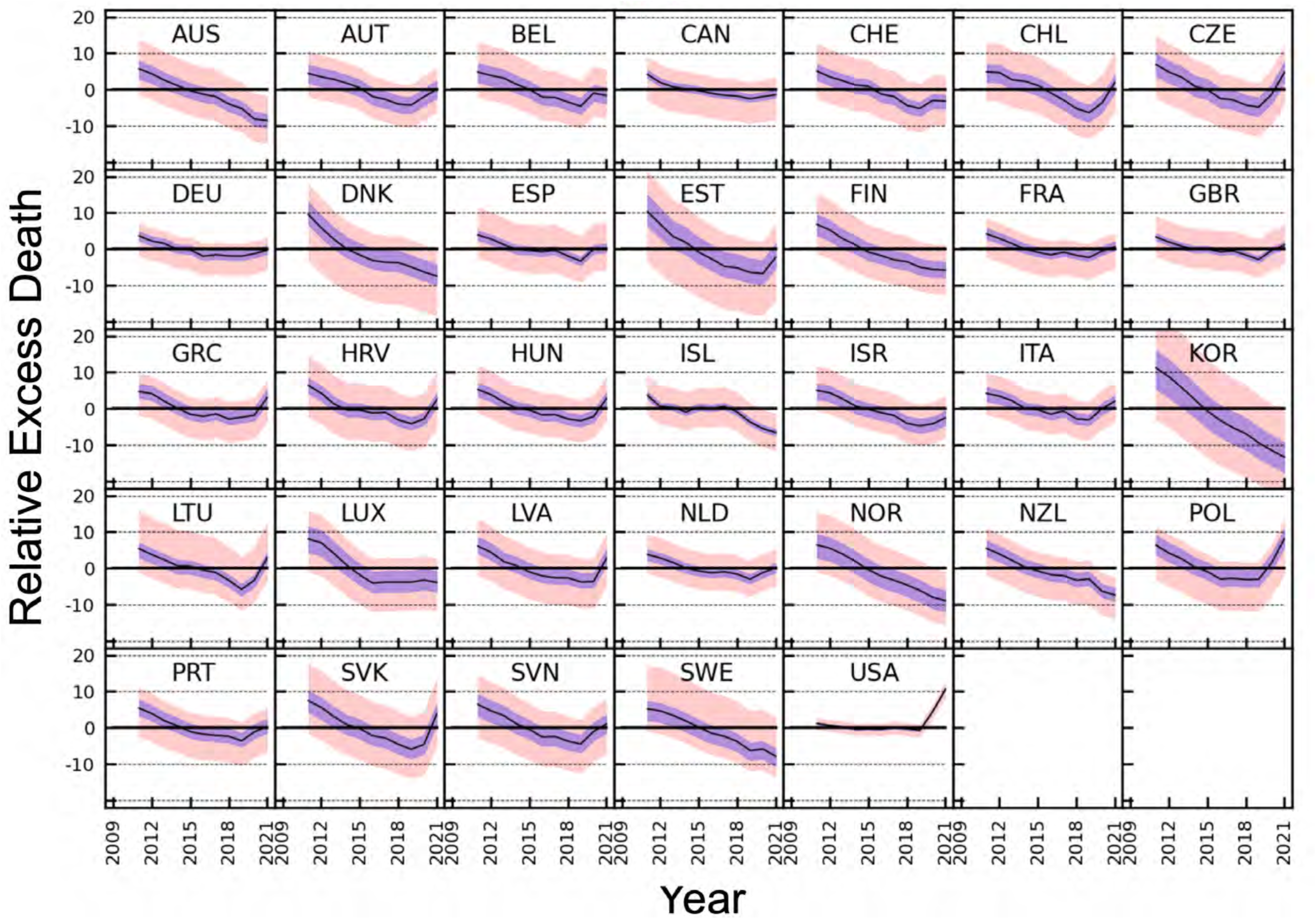
Variation with year from 2009 to 2021 of the Relative Excess Death, p%, averaged over three adjacent years. The salmon shading marks the full range for all 66 reference sets, the purple shading marks the range between the first and third quartile and the black line shows the average over the reference periods. Note that the corresponding figure for averaging over two adjacent years is shown in main text **Figure 2**.

**Figure S4C:**
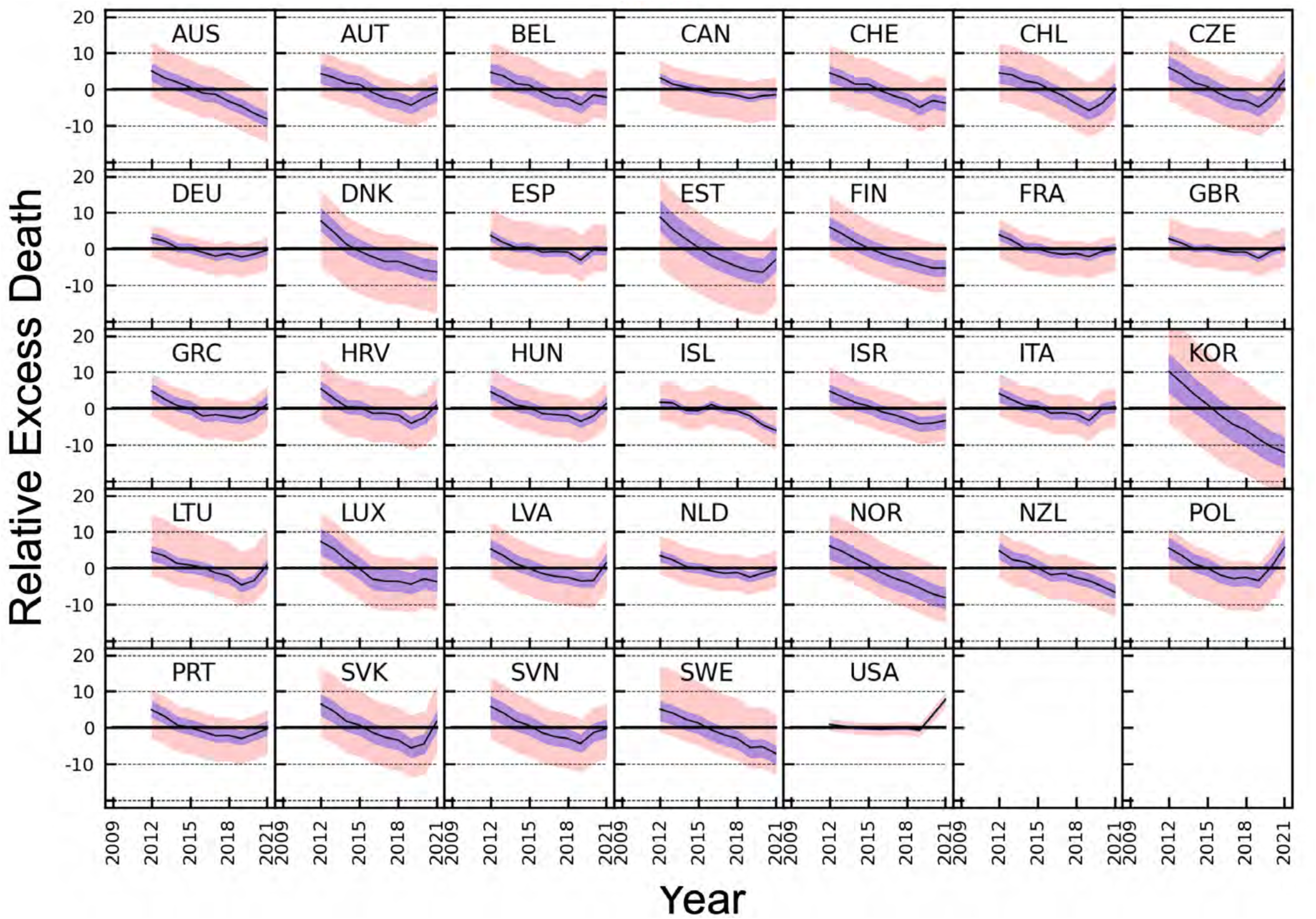
Variation with year from 2009 to 2021 of the relative Excess Death p% averaged over four adjacent years. The salmon shading marks the full range for all 66 reference sets, the purple shading marks the range between the first and third quartile and the black line shows the average over the reference periods.

**Table S1A:** Relative percentage excess death, p%, in a single year for all countries and all years. Countries are listed in alphabetical order. Shading varies from solid green for the lowest values to solid red for the highest values. The shading is calibrated by range of values in all Tables S1 considered together. Note that 2019, the year immediately preceding the pandemics, is ‘greenest’ for all countries (low p%). By the same measure 2009 is ‘reddest’ (high p%).

| Country | 2009 | 2010 | 2011 | 2012 | 2013 | 2014 | 2015 | 2016 | 2017 | 2018 | 2019 | 2020 | 2021 |
| --- | --- | --- | --- | --- | --- | --- | --- | --- | --- | --- | --- | --- | --- |
| Australia | 7.36 | 5.18 | 4.76 | 3.24 | -0.21 | 0.34 | -1.03 | -3.01 | -1.81 | -7.02 | -6.60 | -10.63 | -8.36 |
| Austria | 6.78 | 4.66 | 1.99 | 3.91 | 2.54 | -1.19 | 0.22 | -4.67 | -3.64 | -3.77 | -5.47 | 3.24 | 2.38 |
| Belgium | 7.23 | 5.34 | 2.06 | 4.33 | 3.10 | -3.00 | 0.24 | -3.37 | -3.27 | -3.69 | -6.82 | 7.45 | -5.54 |
| Canada | 7.25 | 3.49 | 2.00 | 0.39 | -0.13 | 0.46 | -1.17 | -2.36 | -1.10 | -1.96 | -4.48 | 0.53 | -0.33 |
| Chile | 4.02 | 8.07 | 2.54 | 3.59 | 1.95 | 1.38 | 0.07 | -3.97 | -4.55 | -7.06 | -7.42 | 3.11 | 9.66 |
| Croatia | 8.98 | 7.17 | 3.09 | 2.63 | -1.70 | -2.15 | 2.41 | -3.84 | -1.63 | -3.87 | -6.86 | 2.03 | 12.04 |
| Czechia | 9.54 | 6.82 | 4.55 | 3.29 | 2.42 | -2.70 | 0.30 | -4.90 | -3.87 | -4.11 | -6.49 | 6.36 | 14.03 |
| Denmark | 13.31 | 10.62 | 4.82 | 2.40 | 0.71 | -2.97 | -2.62 | -3.82 | -4.59 | -2.94 | -7.26 | -8.63 | -6.54 |
| Estonia | 14.52 | 11.13 | 5.58 | 4.07 | 0.77 | 0.67 | -3.98 | -4.77 | -5.58 | -5.26 | -8.49 | -6.51 | 7.99 |
| Finland | 8.21 | 8.01 | 4.32 | 3.62 | 0.64 | -0.73 | -2.40 | -2.21 | -4.33 | -3.89 | -6.80 | -6.08 | -4.54 |
| France | 6.76 | 4.93 | 0.89 | 2.90 | 0.61 | -3.51 | 0.17 | -1.60 | -1.20 | -2.37 | -3.42 | 3.92 | 1.34 |
| Germany | 5.37 | 4.14 | 1.39 | 1.08 | 1.99 | -3.25 | 0.50 | -3.16 | -2.07 | -0.47 | -3.31 | -0.12 | 2.35 |
| Greece | 6.76 | 4.41 | 3.09 | 4.93 | -1.89 | -2.60 | -0.28 | -3.63 | -0.48 | -4.57 | -2.16 | 1.12 | 10.06 |
| Hungary | 6.72 | 5.81 | 3.32 | 2.57 | -0.32 | -1.83 | 0.80 | -4.15 | -1.57 | -2.84 | -5.59 | 1.73 | 11.94 |
| Iceland | 5.23 | 5.20 | 0.67 | -3.92 | 4.30 | -3.34 | 0.26 | 3.13 | -1.54 | -4.32 | -5.16 | -7.05 | -7.56 |
| Israel | 5.89 | 4.83 | 4.43 | 3.86 | -0.17 | -1.31 | 0.69 | -2.72 | -3.55 | -5.83 | -4.78 | -2.09 | -0.93 |
| Italy | 6.11 | 3.08 | 3.50 | 3.67 | -0.60 | -2.85 | 2.23 | -3.95 | -0.26 | -4.22 | -4.84 | 8.86 | 2.10 |
| Latvia | 7.84 | 8.13 | 2.31 | 2.88 | 0.45 | -1.09 | -2.36 | -2.88 | -2.65 | -2.43 | -6.11 | -2.49 | 16.58 |
| Lithuania | 6.54 | 6.64 | 3.06 | 1.60 | 2.24 | -1.71 | 1.24 | -1.06 | -3.71 | -5.16 | -8.61 | 3.90 | 13.35 |
| Luxembourg | 8.10 | 9.30 | 6.89 | 5.08 | 0.37 | -2.75 | -2.97 | -6.44 | -2.11 | -3.04 | -6.21 | -0.48 | -5.04 |
| Netherlands | 5.33 | 4.39 | 1.63 | 2.50 | 0.61 | -2.92 | 0.03 | -0.87 | -1.99 | -2.00 | -4.94 | 3.25 | 1.79 |
| New Zealand | 7.46 | 2.95 | 5.99 | 2.81 | -2.15 | 0.33 | -0.49 | -5.00 | -0.67 | -4.28 | -3.92 | -10.70 | -7.48 |
| Norway | 7.64 | 6.77 | 4.84 | 5.01 | 1.82 | -1.10 | -1.85 | -3.53 | -4.46 | -5.78 | -8.00 | -10.13 | -8.63 |
| Poland | 9.69 | 6.22 | 3.27 | 3.17 | 1.48 | -3.33 | -0.88 | -4.64 | -2.76 | -2.00 | -4.32 | 10.43 | 18.09 |
| Portugal | 8.03 | 7.03 | 1.29 | 3.40 | 0.95 | -2.71 | -1.54 | -1.13 | -3.68 | -2.31 | -4.88 | 3.58 | 2.05 |
| Slovakia | 9.24 | 8.91 | 4.36 | 3.55 | 1.23 | -2.18 | 0.23 | -4.65 | -4.08 | -5.04 | -8.67 | -0.38 | 19.91 |
| Slovenia | 9.46 | 6.01 | 3.94 | 4.07 | 2.06 | -2.80 | -0.66 | -4.14 | -2.45 | -4.56 | -6.44 | 7.54 | 1.86 |
| South Korea | 14.72 | 11.48 | 7.96 | 7.20 | 2.47 | -1.52 | -2.79 | -5.17 | -7.62 | -7.81 | -12.61 | -13.83 | -13.72 |
| Spain | 6.58 | 3.15 | 1.93 | 3.22 | -1.46 | -2.13 | 2.32 | -2.48 | -0.81 | -2.50 | -6.59 | 8.70 | -1.49 |
| Sweden | 6.33 | 5.68 | 3.54 | 4.62 | 1.95 | -0.95 | -0.62 | -3.02 | -3.48 | -4.90 | -10.38 | -2.44 | -10.83 |
| Switzerland | 7.63 | 5.65 | 2.06 | 2.90 | 2.36 | -1.48 | 1.95 | -4.15 | -3.42 | -5.47 | -6.63 | 2.89 | -5.82 |
| United Kingdom | 5.57 | 4.04 | 0.47 | 1.25 | 0.62 | -2.13 | 1.52 | -1.69 | -1.29 | -2.05 | -5.32 | 6.09 | 2.27 |
| United States | 1.67 | 0.89 | 0.92 | 0.01 | -0.20 | -1.18 | 0.30 | -0.50 | 0.43 | -0.92 | -1.69 | 15.73 | 17.65 |

**Table S1B:**
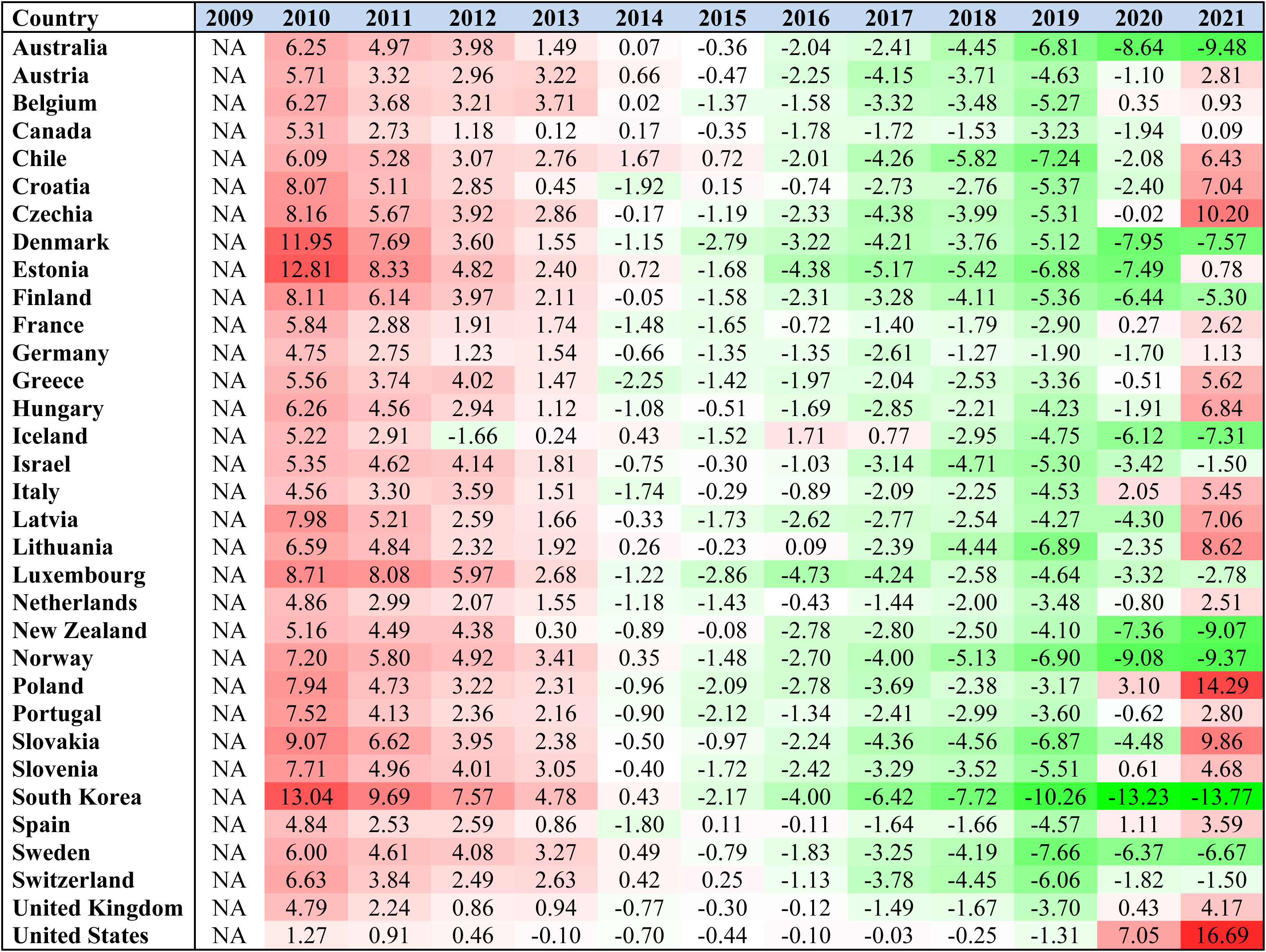
Relative percentage excess death, p%, in two adjacent years for all countries and all years. When considering a two-year projected period, 2009 is added to 2010 and the combined period 2009+2010 is denoted as ‘2010’. For this reason, the entry for 2009 is marked as NA.

**Table S1C:** Relative percentage excess death, p%, in three adjacent years for all countries and all years. When considering three-year projected periods, 2009 and 2010 are added to 2011 and the combined period 2009+2010+2011 is denoted as ‘2011’. For this reason, the entries for both 2009 and 2010 are marked as NA.

| Country | 2009 | 2010 | 2011 | 2012 | 2013 | 2014 | 2015 | 2016 | 2017 | 2018 | 2019 | 2020 | 2021 |
| --- | --- | --- | --- | --- | --- | --- | --- | --- | --- | --- | --- | --- | --- |
| **Australia** | NA | NA | 5.73 | 4.37 | 2.55 | 1.09 | -0.31 | -1.27 | -1.96 | -3.98 | -5.19 | -8.12 | -8.55 |
| **Austria** | NA | NA | 4.45 | 3.52 | 2.82 | 1.72 | 0.51 | -1.91 | -2.72 | -4.02 | -4.30 | -1.98 | 0.07 |
| **Belgium** | NA | NA | 4.84 | 3.90 | 3.17 | 1.43 | 0.10 | -2.05 | -2.15 | -3.45 | -4.61 | -0.98 | -1.63 |
| **Canada** | NA | NA | 4.17 | 1.93 | 0.73 | 0.24 | -0.28 | -1.04 | -1.55 | -1.80 | -2.54 | -1.95 | -1.39 |
| **Chile** | NA | NA | 4.87 | 4.70 | 2.69 | 2.29 | 1.13 | -0.91 | -2.89 | -5.23 | -6.37 | -3.69 | 1.95 |
| **Croatia** | NA | NA | 6.38 | 4.27 | 1.31 | -0.43 | -0.46 | -1.20 | -1.04 | -3.11 | -4.14 | -2.88 | 2.44 |
| **Czechia** | NA | NA | 6.93 | 4.86 | 3.41 | 0.97 | -0.01 | -2.45 | -2.85 | -4.29 | -4.84 | -1.36 | 4.70 |
| **Denmark** | NA | NA | 9.53 | 5.89 | 2.62 | 0.01 | -1.65 | -3.14 | -3.69 | -3.78 | -4.95 | -6.32 | -7.47 |
| **Estonia** | NA | NA | 10.37 | 6.88 | 3.44 | 1.81 | -0.88 | -2.73 | -4.78 | -5.20 | -6.45 | -6.76 | -2.28 |
| **Finland** | NA | NA | 6.81 | 5.28 | 2.83 | 1.14 | -0.86 | -1.79 | -2.99 | -3.49 | -5.02 | -5.60 | -5.79 |
| **France** | NA | NA | 4.15 | 2.89 | 1.47 | -0.05 | -0.91 | -1.63 | -0.88 | -1.73 | -2.34 | -0.60 | 0.63 |
| **Germany** | NA | NA | 3.61 | 2.18 | 1.49 | -0.09 | -0.26 | -1.97 | -1.59 | -1.89 | -1.96 | -1.30 | -0.33 |
| **Greece** | NA | NA | 4.70 | 4.15 | 2.00 | 0.08 | -1.57 | -2.17 | -1.46 | -2.89 | -2.41 | -1.85 | 3.06 |
| **Hungary** | NA | NA | 5.27 | 3.89 | 1.84 | 0.12 | -0.44 | -1.74 | -1.65 | -2.85 | -3.35 | -2.22 | 2.73 |
| **Iceland** | NA | NA | 3.67 | 0.58 | 0.38 | -0.98 | 0.38 | 0.07 | 0.60 | -0.97 | -3.71 | -5.53 | -6.61 |
| **Israel** | NA | NA | 5.03 | 4.36 | 2.65 | 0.74 | -0.26 | -1.12 | -1.89 | -4.06 | -4.73 | -4.20 | -2.56 |
| **Italy** | NA | NA | 4.19 | 3.42 | 2.16 | 0.02 | -0.39 | -1.53 | -0.68 | -2.81 | -3.12 | -0.01 | 2.07 |
| **Latvia** | NA | NA | 6.08 | 4.43 | 1.87 | 0.73 | -1.01 | -2.12 | -2.63 | -2.65 | -3.73 | -3.68 | 2.68 |
| **Lithuania** | NA | NA | 5.41 | 3.75 | 2.30 | 0.70 | 0.59 | -0.50 | -1.18 | -3.31 | -5.83 | -3.28 | 2.89 |
| **Luxembourg** | NA | NA | 8.09 | 7.05 | 4.04 | 0.80 | -1.83 | -4.09 | -3.83 | -3.83 | -3.82 | -3.23 | -3.90 |
| **Netherlands** | NA | NA | 3.75 | 2.83 | 1.57 | 0.02 | -0.77 | -1.24 | -0.96 | -1.63 | -3.00 | -1.19 | 0.08 |
| **New Zealand** | NA | NA | 5.45 | 3.92 | 2.15 | 0.31 | -0.75 | -1.77 | -2.05 | -3.31 | -2.98 | -6.36 | -7.40 |
| **Norway** | NA | NA | 6.40 | 5.53 | 3.88 | 1.88 | -0.39 | -2.17 | -3.29 | -4.60 | -6.10 | -8.00 | -8.93 |
| **Poland** | NA | NA | 6.35 | 4.20 | 2.62 | 0.39 | -0.93 | -2.96 | -2.78 | -3.12 | -3.03 | 1.43 | 8.18 |
| **Portugal** | NA | NA | 5.39 | 3.88 | 1.88 | 0.50 | -1.12 | -1.79 | -2.13 | -2.38 | -3.63 | -1.17 | 0.29 |
| **Slovakia** | NA | NA | 7.48 | 5.58 | 3.03 | 0.83 | -0.25 | -2.22 | -2.86 | -4.59 | -5.96 | -4.67 | 3.80 |
| **Slovenia** | NA | NA | 6.42 | 4.66 | 3.34 | 1.05 | -0.49 | -2.55 | -2.43 | -3.72 | -4.52 | -1.08 | 1.04 |
| **South Korea** | NA | NA | 11.26 | 8.83 | 5.80 | 2.59 | -0.69 | -3.21 | -5.26 | -6.91 | -9.42 | -11.50 | -13.40 |
| **Spain** | NA | NA | 3.84 | 2.77 | 1.21 | -0.16 | -0.40 | -0.77 | -0.35 | -1.93 | -3.34 | -0.07 | 0.24 |
| **Sweden** | NA | NA | 5.17 | 4.61 | 3.36 | 1.85 | 0.11 | -1.54 | -2.39 | -3.81 | -6.29 | -5.89 | -7.89 |
| **Switzerland** | NA | NA | 5.07 | 3.52 | 2.44 | 1.23 | 0.94 | -1.25 | -1.91 | -4.35 | -5.20 | -3.01 | -3.18 |
| **United Kingdom** | NA | NA | 3.31 | 1.90 | 0.78 | -0.11 | 0.00 | -0.78 | -0.52 | -1.68 | -2.91 | -0.39 | 1.05 |
| **United States** | NA | NA | 1.15 | 0.60 | 0.24 | -0.47 | -0.36 | -0.46 | 0.08 | -0.33 | -0.74 | 4.42 | 10.60 |

**Table S1D:**
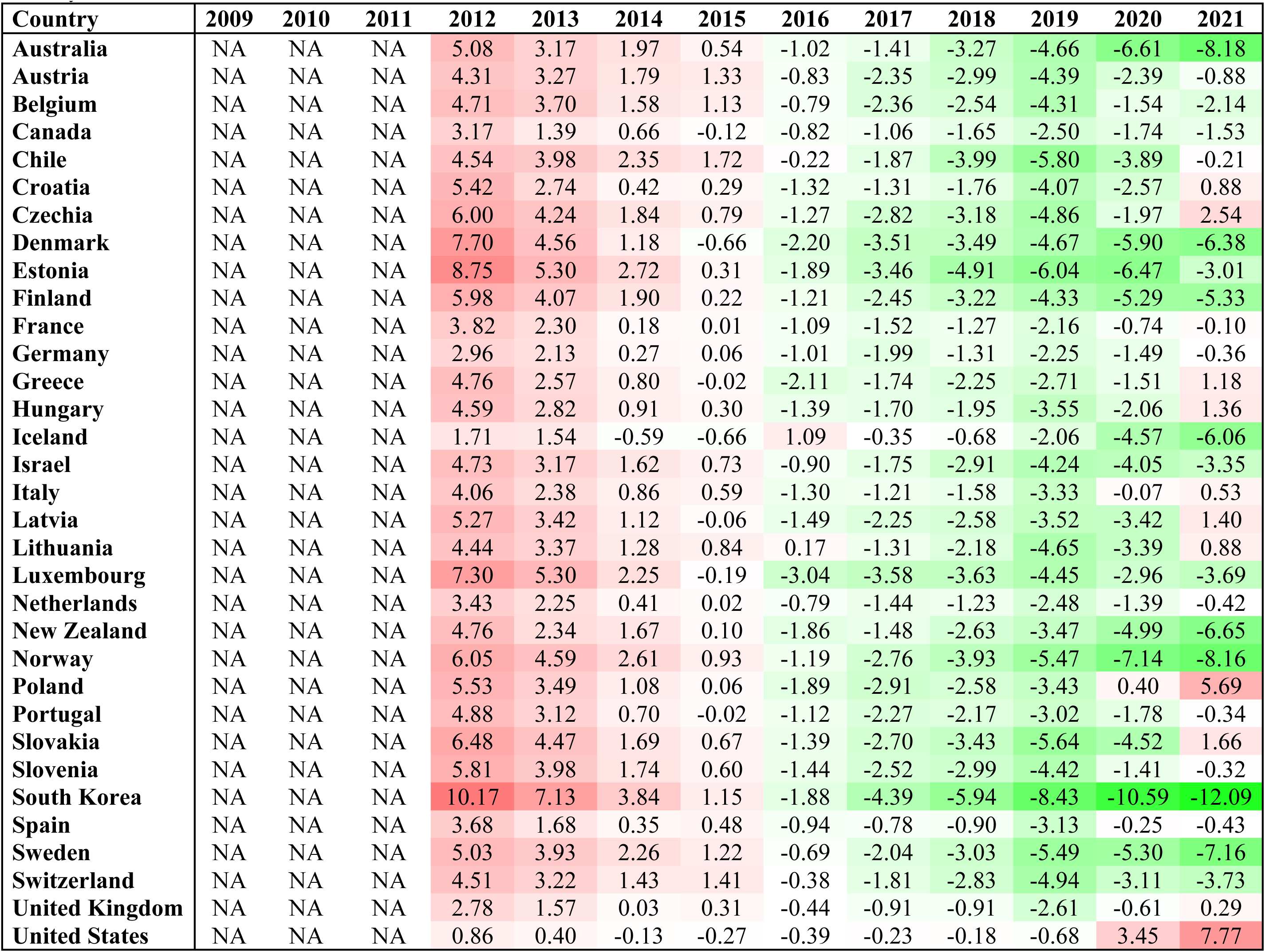
Percentage excess death, p%, in four adjacent years for all countries and all years. When considering four-year projected periods, 2009, 2010 and 2011 are added to 2012 and the combined period 2009+2010+2011+2012 is denoted as ‘2012’. For this reason, the entries for both 2009, 2010 and 2011 are marked as NA. With a longer projected period, the relative percentage excess death gets smaller for every country.

| Country | 2009 | 2010 | 2011 | 2012 | 2013 | 2014 | 2015 | 2016 | 2017 | 2018 | 2019 | 2020 | 2021 |
| --- | --- | --- | --- | --- | --- | --- | --- | --- | --- | --- | --- | --- | --- |
| Australia | NA | NA | NA | 5.08 | 3.17 | 1.97 | 0.54 | -1.02 | -1.41 | -3.27 | -4.66 | -6.61 | -8.18 |
| Austria | NA | NA | NA | 4.31 | 3.27 | 1.79 | 1.33 | -0.83 | -2.35 | -2.99 | -4.39 | -2.39 | -0.88 |
| Belgium | NA | NA | NA | 4.71 | 3.70 | 1.58 | 1.13 | -0.79 | -2.36 | -2.54 | -4.31 | -1.54 | -2.14 |
| Canada | NA | NA | NA | 3.17 | 1.39 | 0.66 | -0.12 | -0.82 | -1.06 | -1.65 | -2.50 | -1.74 | -1.53 |
| Chile | NA | NA | NA | 4.54 | 3.98 | 2.35 | 1.72 | -0.22 | -1.87 | -3.99 | -5.80 | -3.89 | -0.21 |
| Croatia | NA | NA | NA | 5.42 | 2.74 | 0.42 | 0.29 | -1.32 | -1.31 | -1.76 | -4.07 | -2.57 | 0.88 |
| Czechia | NA | NA | NA | 6.00 | 4.24 | 1.84 | 0.79 | -1.27 | -2.82 | -3.18 | -4.86 | -1.97 | 2.54 |
| Denmark | NA | NA | NA | 7.70 | 4.56 | 1.18 | -0.66 | -2.20 | -3.51 | -3.49 | -4.67 | -5.90 | -6.38 |
| Estonia | NA | NA | NA | 8.75 | 5.30 | 2.72 | 0.31 | -1.89 | -3.46 | -4.91 | -6.04 | -6.47 | -3.01 |
| Finland | NA | NA | NA | 5.98 | 4.07 | 1.90 | 0.22 | -1.21 | -2.45 | -3.22 | -4.33 | -5.29 | -5.33 |
| France | NA | NA | NA | 3.82 | 2.30 | 0.18 | 0.01 | -1.09 | -1.52 | -1.27 | -2.16 | -0.74 | -0.10 |
| Germany | NA | NA | NA | 2.96 | 2.13 | 0.27 | 0.06 | -1.01 | -1.99 | -1.31 | -2.25 | -1.49 | -0.36 |
| Greece | NA | NA | NA | 4.76 | 2.57 | 0.80 | -0.02 | -2.11 | -1.74 | -2.25 | -2.71 | -1.51 | 1.18 |
| Hungary | NA | NA | NA | 4.59 | 2.82 | 0.91 | 0.30 | -1.39 | -1.70 | -1.95 | -3.55 | -2.06 | 1.36 |
| Iceland | NA | NA | NA | 1.71 | 1.54 | -0.59 | -0.66 | 1.09 | -0.35 | -0.68 | -2.06 | -4.57 | -6.06 |
| Israel | NA | NA | NA | 4.73 | 3.17 | 1.62 | 0.73 | -0.90 | -1.75 | -2.91 | -4.24 | -4.05 | -3.35 |
| Italy | NA | NA | NA | 4.06 | 2.38 | 0.86 | 0.59 | -1.30 | -1.21 | -1.58 | -3.33 | -0.07 | 0.53 |
| Latvia | NA | NA | NA | 5.27 | 3.42 | 1.12 | -0.06 | -1.49 | -2.25 | -2.58 | -3.52 | -3.42 | 1.40 |
| Lithuania | NA | NA | NA | 4.44 | 3.37 | 1.28 | 0.84 | 0.17 | -1.31 | -2.18 | -4.65 | -3.39 | 0.88 |
| Luxembourg | NA | NA | NA | 7.30 | 5.30 | 2.25 | -0.19 | -3.04 | -3.58 | -3.63 | -4.45 | -2.96 | -3.69 |
| Netherlands | NA | NA | NA | 3.43 | 2.25 | 0.41 | 0.02 | -0.79 | -1.44 | -1.23 | -2.48 | -1.39 | -0.42 |
| New Zealand | NA | NA | NA | 4.76 | 2.34 | 1.67 | 0.10 | -1.86 | -1.48 | -2.63 | -3.47 | -4.99 | -6.65 |
| Norway | NA | NA | NA | 6.05 | 4.59 | 2.61 | 0.93 | -1.19 | -2.76 | -3.93 | -5.47 | -7.14 | -8.16 |
| Poland | NA | NA | NA | 5.53 | 3.49 | 1.08 | 0.06 | -1.89 | -2.91 | -2.58 | -3.43 | 0.40 | 5.69 |
| Portugal | NA | NA | NA | 4.88 | 3.12 | 0.70 | -0.02 | -1.12 | -2.27 | -2.17 | -3.02 | -1.78 | -0.34 |
| Slovakia | NA | NA | NA | 6.48 | 4.47 | 1.69 | 0.67 | -1.39 | -2.70 | -3.43 | -5.64 | -4.52 | 1.66 |
| Slovenia | NA | NA | NA | 5.81 | 3.98 | 1.74 | 0.60 | -1.44 | -2.52 | -2.99 | -4.42 | -1.41 | -0.32 |
| South Korea | NA | NA | NA | 10.17 | 7.13 | 3.84 | 1.15 | -1.88 | -4.39 | -5.94 | -8.43 | -10.59 | -12.09 |
| Spain | NA | NA | NA | 3.68 | 1.68 | 0.35 | 0.48 | -0.94 | -0.78 | -0.90 | -3.13 | -0.25 | -0.43 |
| Sweden | NA | NA | NA | 5.03 | 3.93 | 2.26 | 1.22 | -0.69 | -2.04 | -3.03 | -5.49 | -5.30 | -7.16 |
| Switzerland | NA | NA | NA | 4.51 | 3.22 | 1.43 | 1.41 | -0.38 | -1.81 | -2.83 | -4.94 | -3.11 | -3.73 |
| United Kingdom | NA | NA | NA | 2.78 | 1.57 | 0.03 | 0.31 | -0.44 | -0.91 | -0.91 | -2.61 | -0.61 | 0.29 |
| United States | NA | NA | NA | 0.86 | 0.40 | -0.13 | -0.27 | -0.39 | -0.23 | -0.18 | -0.68 | 3.45 | 7.77 |

**Table S2:** The percentage excess death , p%, of the worst, the second worst and third worst pairs of adjacent years compared to the corresponding value in two-year pandemic projected period 2020+2021. Those entries where the two-year pandemic period 2020+2021 are worst are shown in bold type.

| Country | Worst<br>Pair Years | 2nd Worst<br>Pair Years | 3rd Worst<br>Pair Years | Worst<br>p% | 2nd Worst<br>p% | 3rd Worst<br>p% | 2020+2021<br>p% | Worst minus<br>(2020+2021) p% |
| --- | --- | --- | --- | --- | --- | --- | --- | --- |
| Australia | 2009+2010 | 2010+2011 | 2011+2012 | 6.2 | 5.0 | 4.0 | -9.5 | 15.7 |
| Austria | 2009+2010 | 2010+2011 | 2012+2013 | 5.7 | 3.3 | 3.2 | 2.8 | 2.9 |
| Belgium | 2009+2010 | 2012+2013 | 2010+2011 | 6.3 | 3.7 | 3.7 | 0.9 | 5.3 |
| Canada | 2009+2010 | 2010+2011 | 2011+2012 | 5.3 | 2.7 | 1.2 | 0.1 | 5.2 |
| Switzerland | 2009+2010 | 2010+2011 | 2012+2013 | 6.6 | 3.8 | 2.6 | -1.5 | 8.1 |
| Chile | **2020+2021** | 2009+2010 | 2010+2011 | **6.4** | 6.1 | 5.3 | 6.4 | 0.0 |
| Czechia | **2020+2021** | 2009+2010 | 2010+2011 | **10.2** | 8.2 | 5.7 | 10.2 | 0.0 |
| Germany | 2009+2010 | 2010+2011 | 2012+2013 | 4.8 | 2.8 | 1.5 | 1.1 | 3.6 |
| Denmark | 2009+2010 | 2010+2011 | 2011+2012 | 12.0 | 7.7 | 3.6 | -7.6 | 19.5 |
| Spain | 2009+2010 | **2020+2021** | 2011+2012 | 4.8 | **3.6** | 2.6 | 3.6 | 1.3 |
| Estonia | 2009+2010 | 2010+2011 | 2011+2012 | 12.8 | 8.3 | 4.8 | 0.8 | 12.0 |
| Finland | 2009+2010 | 2010+2011 | 2011+2012 | 8.1 | 6.1 | 4.0 | -5.3 | 13.4 |
| France | 2009+2010 | 2010+2011 | **2020+2021** | 5.8 | 2.9 | **2.6** | 2.6 | 3.2 |
| United Kingdom | 2009+2010 | **2020+2021** | 2010+2011 | 4.8 | **4.2** | 2.2 | 4.2 | 0.6 |
| Greece | **2020+2021** | 2009+2010 | 2011+2012 | **5.6** | 5.6 | 4.0 | 5.6 | 0.0 |
| Croatia | 2009+2010 | **2020+2021** | 2010+2011 | 8.1 | **7.0** | 5.1 | 7.0 | 1.0 |
| Hungary | **2020+2021** | 2009+2010 | 2010+2011 | **6.8** | 6.3 | 4.6 | 6.8 | 0.0 |
| Iceland | 2009+2010 | 2010+2011 | 2015+2016 | 5.2 | 2.9 | 1.7 | -7.3 | 12.5 |
| Israel | 2009+2010 | 2010+2011 | 2011+2012 | 5.4 | 4.6 | 4.1 | -1.5 | 6.9 |
| Italy | **2020+2021** | 2009+2010 | 2011+2012 | **5.5** | 4.6 | 3.6 | 5.5 | 0.0 |
| South Korea | 2009+2010 | 2010+2011 | 2011+2012 | 13.0 | 9.7 | 7.6 | -13.8 | 26.8 |
| Lithuania | **2020+2021** | 2009+2010 | 2010+2011 | **8.6** | 6.6 | 4.8 | 8.6 | 0.0 |
| Luxembourg | 2009+2010 | 2010+2011 | 2011+2012 | 8.7 | 8.1 | 6.0 | -2.8 | 11.5 |
| Latvia | 2009+2010 | **2020+2021** | 2010+2011 | 8.0 | **7.1** | 5.2 | 7.1 | 0.9 |
| Netherlands | 2009+2010 | 2010+2011 | **2020+2021** | 4.9 | 3.0 | **2.5** | 2.5 | 2.3 |
| Norway | 2009+2010 | 2010+2011 | 2011+2012 | 7.2 | 5.8 | 4.9 | -9.4 | 16.6 |
| New Zealand | 2009+2010 | 2010+2011 | 2011+2012 | 5.2 | 4.5 | 4.4 | -9.1 | 14.2 |
| Poland | **2020+2021** | 2009+2010 | 2010+2011 | **14.3** | 7.9 | 4.7 | 14.3 | 0.0 |
| Portugal | 2009+2010 | 2010+2011 | **2020+2021** | 7.5 | 4.1 | **2.8** | 2.8 | 4.7 |
| Slovakia | **2020+2021** | 2009+2010 | 2010+2011 | 9.9 | 9.1 | 6.6 | 9.9 | 0.0 |
| Slovenia | 2009+2010 | 2010+2011 | **2020+2021** | **7.7** | 5.0 | **4.7** | 4.7 | 3.0 |
| Sweden | 2009+2010 | 2010+2011 | 2011+2012 | 6.0 | 4.6 | 4.1 | -6.7 | 12.7 |
| United States | **2020+2021** | 2019+2020 | 2009+2010 | **16.7** | 7.0 | 1.3 | 16.7 | 0.0 |

**Table S3:**
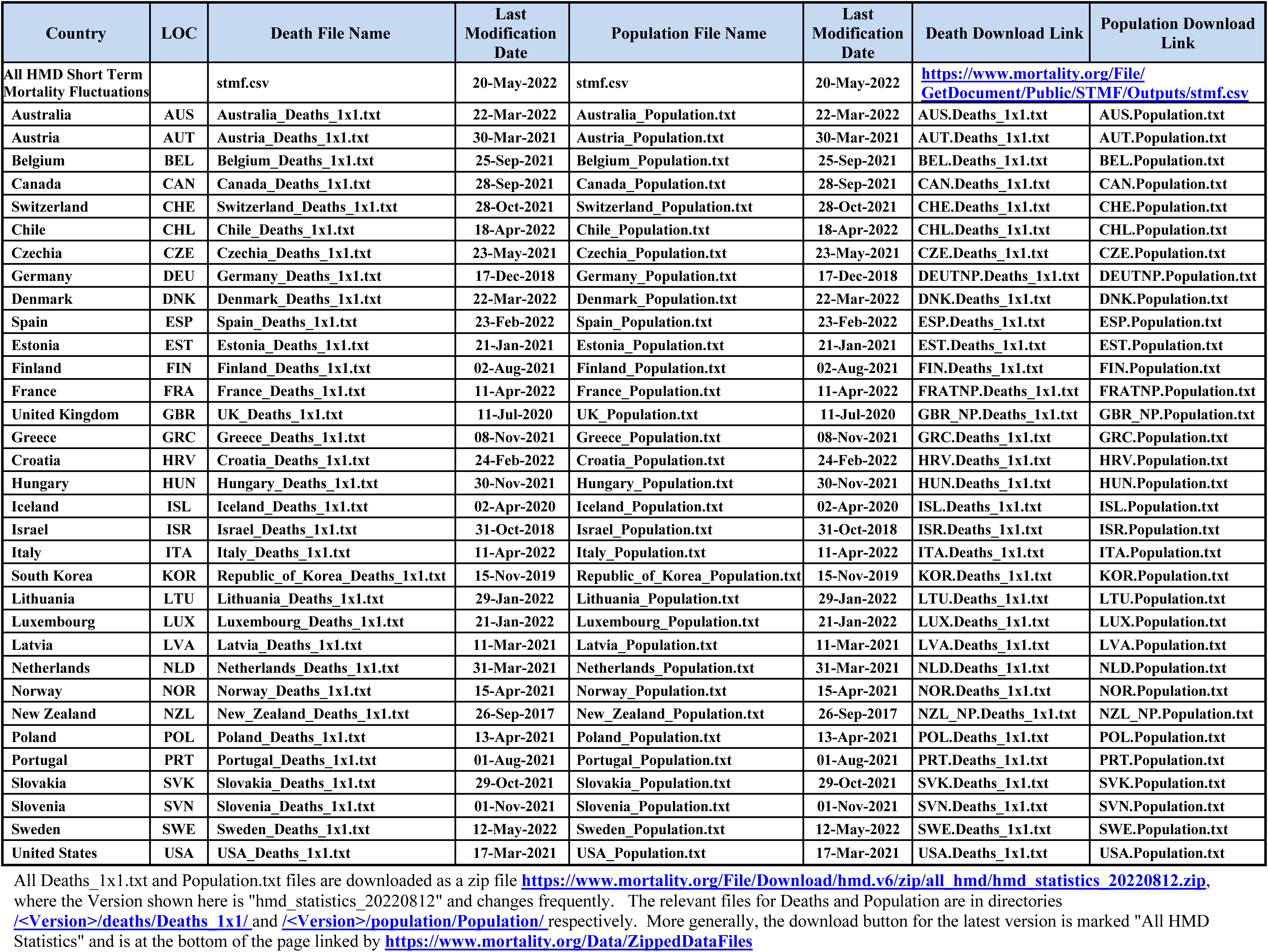
Supplementary links to data sources.

| Country | LOC | Death File Name | Last Modification Date | Population File Name | Last Modification Date | Death Download Link | Population Download Link |
| --- | --- | --- | --- | --- | --- | --- | --- |
| All HMD Short Term Mortality Fluctuations |  | stmf.csv | 20-May-2022 | stmf.csv | 20-May-2022 | https://www.mortality.org/File/GetDocument/Public/STMF/Outputs/stmf.csv |  |
| Australia | AUS | Australia_Deaths_1x1.txt | 22-Mar-2022 | Australia_Population.txt | 22-Mar-2022 | AUS.Deaths_1x1.txt | AUS.Population.txt |
| Austria | AUT | Austria_Deaths_1x1.txt | 30-Mar-2021 | Austria_Population.txt | 30-Mar-2021 | AUT.Deaths_1x1.txt | AUT.Population.txt |
| Belgium | BEL | Belgium_Deaths_1x1.txt | 25-Sep-2021 | Belgium_Population.txt | 25-Sep-2021 | BEL.Deaths_1x1.txt | BEL.Population.txt |
| Canada | CAN | Canada_Deaths_1x1.txt | 28-Sep-2021 | Canada_Population.txt | 28-Sep-2021 | CAN.Deaths_1x1.txt | CAN.Population.txt |
| Switzerland | CHE | Switzerland_Deaths_1x1.txt | 28-Oct-2021 | Switzerland_Population.txt | 28-Oct-2021 | CHE.Deaths_1x1.txt | CHE.Population.txt |
| Chile | CHL | Chile_Deaths_1x1.txt | 18-Apr-2022 | Chile_Population.txt | 18-Apr-2022 | CHL.Deaths_1x1.txt | CHL.Population.txt |
| Czechia | CZE | Czechia_Deaths_1x1.txt | 23-May-2021 | Czechia_Population.txt | 23-May-2021 | CZE.Deaths_1x1.txt | CZE.Population.txt |
| Germany | DEU | Germany_Deaths_1x1.txt | 17-Dec-2018 | Germany_Population.txt | 17-Dec-2018 | DEUTNP.Deaths_1x1.txt | DEUTNP.Population.txt |
| Denmark | DNK | Denmark_Deaths_1x1.txt | 22-Mar-2022 | Denmark_Population.txt | 22-Mar-2022 | DNK.Deaths_1x1.txt | DNK.Population.txt |
| Spain | ESP | Spain_Deaths_1x1.txt | 23-Feb-2022 | Spain_Population.txt | 23-Feb-2022 | ESP.Deaths_1x1.txt | ESP.Population.txt |
| Estonia | EST | Estonia_Deaths_1x1.txt | 21-Jan-2021 | Estonia_Population.txt | 21-Jan-2021 | EST.Deaths_1x1.txt | EST.Population.txt |
| Finland | FIN | Finland_Deaths_1x1.txt | 02-Aug-2021 | Finland_Population.txt | 02-Aug-2021 | FIN.Deaths_1x1.txt | FIN.Population.txt |
| France | FRA | France_Deaths_1x1.txt | 11-Apr-2022 | France_Population.txt | 11-Apr-2022 | FRATNP.Deaths_1x1.txt | FRATNP.Population.txt |
| United Kingdom | GBR | UK_Deaths_1x1.txt | 11-Jul-2020 | UK_Population.txt | 11-Jul-2020 | GBR_NP.Deaths_1x1.txt | GBR_NP.Population.txt |
| Greece | GRC | Greece_Deaths_1x1.txt | 08-Nov-2021 | Greece_Population.txt | 08-Nov-2021 | GRC.Deaths_1x1.txt | GRC.Population.txt |
| Croatia | HRV | Croatia_Deaths_1x1.txt | 24-Feb-2022 | Croatia_Population.txt | 24-Feb-2022 | HRV.Deaths_1x1.txt | HRV.Population.txt |
| Hungary | HUN | Hungary_Deaths_1x1.txt | 30-Nov-2021 | Hungary_Population.txt | 30-Nov-2021 | HUN.Deaths_1x1.txt | HUN.Population.txt |
| Iceland | ISL | Iceland_Deaths_1x1.txt | 02-Apr-2020 | Iceland_Population.txt | 02-Apr-2020 | ISL.Deaths_1x1.txt | ISL.Population.txt |
| Israel | ISR | Israel_Deaths_1x1.txt | 31-Oct-2018 | Israel_Population.txt | 31-Oct-2018 | ISR.Deaths_1x1.txt | ISR.Population.txt |
| Italy | ITA | Italy_Deaths_1x1.txt | 11-Apr-2022 | Italy_Population.txt | 11-Apr-2022 | ITA.Deaths_1x1.txt | ITA.Population.txt |
| South Korea | KOR | Republic_of_Korea_Deaths_1x1.txt | 15-Nov-2019 | Republic_of_Korea_Population.txt | 15-Nov-2019 | KOR.Deaths_1x1.txt | KOR.Population.txt |
| Lithuania | LTU | Lithuania_Deaths_1x1.txt | 29-Jan-2022 | Lithuania_Population.txt | 29-Jan-2022 | LTU.Deaths_1x1.txt | LTU.Population.txt |
| Luxembourg | LUX | Luxembourg_Deaths_1x1.txt | 21-Jan-2022 | Luxembourg_Population.txt | 21-Jan-2022 | LUX.Deaths_1x1.txt | LUX.Population.txt |
| Latvia | LVA | Latvia_Deaths_1x1.txt | 11-Mar-2021 | Latvia_Population.txt | 11-Mar-2021 | LVA.Deaths_1x1.txt | LVA.Population.txt |
| Netherlands | NLD | Netherlands_Deaths_1x1.txt | 31-Mar-2021 | Netherlands_Population.txt | 31-Mar-2021 | NLD.Deaths_1x1.txt | NLD.Population.txt |
| Norway | NOR | Norway_Deaths_1x1.txt | 15-Apr-2021 | Norway_Population.txt | 15-Apr-2021 | NOR.Deaths_1x1.txt | NOR.Population.txt |
| New Zealand | NZL | New_Zealand_Deaths_1x1.txt | 26-Sep-2017 | New_Zealand_Population.txt | 26-Sep-2017 | NZL_NP.Deaths_1x1.txt | NZL_NP.Population.txt |
| Poland | POL | Poland_Deaths_1x1.txt | 13-Apr-2021 | Poland_Population.txt | 13-Apr-2021 | POL.Deaths_1x1.txt | POL.Population.txt |
| Portugal | PRT | Portugal_Deaths_1x1.txt | 01-Aug-2021 | Portugal_Population.txt | 01-Aug-2021 | PRT.Deaths_1x1.txt | PRT.Population.txt |
| Slovakia | SVK | Slovakia_Deaths_1x1.txt | 29-Oct-2021 | Slovakia_Population.txt | 29-Oct-2021 | SVK.Deaths_1x1.txt | SVK.Population.txt |
| Slovenia | SVN | Slovenia_Deaths_1x1.txt | 01-Nov-2021 | Slovenia_Population.txt | 01-Nov-2021 | SVN.Deaths_1x1.txt | SVN.Population.txt |
| Sweden | SWE | Sweden_Deaths_1x1.txt | 12-May-2022 | Sweden_Population.txt | 12-May-2022 | SWE.Deaths_1x1.txt | SWE.Population.txt |
| United States | USA | USA_Deaths_1x1.txt | 17-Mar-2021 | USA_Population.txt | 17-Mar-2021 | USA.Deaths_1x1.txt | USA.Population.txt |

All Deaths\_1x1.txt and Population.txt files are downloaded as a zip file https://www.mortality.org/File/Download/hmd.v6/zip/all_hmd/hmd_statistics_20220812.zip, where the Version shown here is "hmd\_statistics\_20220812" and changes frequently. The relevant files for Deaths and Population are in directories [<Version>/deaths/Deaths\_1x1/](https://www.mortality.org/File/Download/hmd.v6/zip/all_hmd/hmd_statistics_20220812.zip/deaths/Deaths_1x1/) and [<Version>/population/Population/](https://www.mortality.org/File/Download/hmd.v6/zip/all_hmd/hmd_statistics_20220812.zip/population/Population/) respectively. More generally, the download button for the latest version is marked "All HMD Statistics" and is at the bottom of the page linked by https://www.mortality.org/Data/ZippedDataFiles

## REFERENCES

1. Kiang MV, Irizarry RA, Buckee CO, Balsari S. Every Body Counts: Measuring Mortality From the COVID-19 Pandemic. Ann Intern Med. 2020 Dec 15;173(12):1004–1007.

2. Islam N. “Excess deaths" is the best metric for tracking the pandemic. BMJ. 2022 Feb 4;376:o285.

3. Vandenbroucke JP. Covid-19: excess deaths should be the outcome measure. Ned Tijdschr Geneeskd. 2021 Sep 7;165:D6219.

4. COVID-19 Excess Mortality Collaborators. Estimating excess mortality due to the COVID-19 pandemic: a systematic analysis of COVID-19-related mortality, 2020–21. Lancet, March 10, 2022 DOI:https://doi.org/10.1016/S0140-6736(21)02796-3

5. World Health Organization, Global excess deaths associated with COVID-19, January 2020 - December 2021. In: https://www.who.int/data/stories/global-excess-deaths-associated-with-covid-19-january-2020-december-2021, last accessed May 6, 2022.

6. Levitt M, Zonta F, Ioannidis JPA. Comparison of pandemic excess mortality in 2020-2021 across different empirical calculations. Env Res 2022; 213:113754.

7. Nepomuceno MR, Klimkin I, Jdanov DA, Aluztiza-Galarza A, Shkolnikov VM. Sensitivity analysis of excess mortality due to the COVID-19 pandemic. Population and Development Review 2022, first published: 03 March 2022, https://doi.org/10.1111/padr.12475.

8. Kowall B, Standl F, Oesterling F, Brune B, Brinkmann M, Dudda M, Pflaumer P, Jöckel KH, Stang A. Excess mortality due to Covid-19? A comparison of total mortality in 2020 with total mortality in 2016 to 2019 in Germany, Sweden and Spain. PLoS One. 2021 Aug 3;16(8):e0255540.

9. Gianicolo EAL, Russo A, Büchler B, Taylor K, Stang A, Blettner M. Gender specific excess mortality in Italy during the COVID-19 pandemic accounting for age. Eur J Epidemiol. 2021 Feb;36(2):213–218.

10. >Schöley J. Robustness and bias of European excess death estimates in 2020 under varying model specifications. medRxiv 2021.06.04.21258353.

11. Dafflon J, F Da Costa P, Váša F, Monti RP, Bzdok D, Hellyer PJ, Turkheimer F, Smallwood J, Jones E, Leech R. A guided multiverse study of neuroimaging analyses. Nat Commun. 2022 Jun 29;13(1):3758.

12. Weermeijer J, Lafit G, Kiekens G, Wampers M, Eisele G, Kasanova Z, Vaessen T, Kuppens P, Myin-Germeys I. Applying multiverse analysis to experience sampling data: Investigating whether preprocessing choices affect robustness of conclusions. Behav Res Methods. 2022 Feb 9. doi: 10.3758/s13428-021-01777-1.

13. Kołodziej A, Magnuski M, Ruban A, Brzezicka A. No relationship between frontal alpha asymmetry and depressive disorders in a multiverse analysis of five studies. Elife. 2021 May 26;10:e60595.

14. Steegen S, Tuerlinckx F, Gelman A, Vanpaemel W. Increasing Transparency Through a Multiverse Analysis. Perspect Psychol Sci. 2016 Sep;11(5):702–712.

15. Sala-i-Martin XX. I just run four million regressions. National Bureau of Economic Research, working paper 6252. Accessible in www.nber.org/paper/w6252.

16. Klau S, Hoffmann S, Patel CJ, Ioannidis JP, Boulesteix AL. Examining the robustness of observational associations to model, measurement and sampling uncertainty with the vibration of effects framework. Int J Epidemiol. 2021 Mar 3;50(1):266–278.

17. Patel CJ, Burford B, Ioannidis JP. Assessment of vibration of effects due to model specification can demonstrate the instability of observational associations. J Clin Epidemiol. 2015 Sep;68(9):1046–58.

18. Ioannidis JP. Why most discovered true associations are inflated. Epidemiology. 2008 Sep;19(5):640–8.

19. Aczel B, Szaszi B, Nilsonne G, van den Akker OR, Albers CJ, van Assen MA, Bastiaansen JA, Benjamin D, Boehm U, Botvinik-Nezer R, Bringmann LF, Busch NA, Caruyer E, Cataldo AM, Cowan N, Delios A, van Dongen NN, Donkin C, van Doorn JB, Dreber A, Dutilh G, Egan GF, Gernsbacher MA, Hoekstra R, Hoffmann S, Holzmeister F, Huber J, Johannesson M, Jonas KJ, Kindel AT, Kirchler M, Kunkels YK, Lindsay DS, Mangin JF, Matzke D, Munafò MR, Newell BR, Nosek BA, Poldrack RA, van Ravenzwaaij D, Rieskamp J, Salganik MJ, Sarafoglou A, Schonberg T, Schweinsberg M, Shanks D, Silberzahn R, Simons DJ, Spellman BA, St-Jean S, Starns JJ, Uhlmann EL, Wicherts J, Wagenmakers EJ. 1. Consensus-based guidance for conducting and reporting multi-analyst studies. Elife. 2021 Nov 9;10:e72185.

20. Botvinik-Nezer R, Holzmeister F, Camerer CF, Dreber A, Huber J, Johannesson M, Kirchler M, Iwanir R, Mumford JA, Adcock RA, Avesani P, Baczkowski BM, Bajracharya A, Bakst L, Ball S, Barilari M, Bault N, Beaton D, Beitner J, Benoit RG, Berkers RMWJ, Bhanji JP, Biswal BB, Bobadilla-Suarez S, Bortolini T, Bottenhorn KL, Bowring A, Braem S, Brooks HR, Brudner EG, Calderon CB, Camilleri JA, Castrellon JJ, Cecchetti L, Cieslik EC, Cole ZJ, Collignon O, Cox RW, Cunningham WA, Czoschke S, Dadi K, Davis CP, Luca A, Delgado MR, Demetriou L, Dennison JB, Di X, Dickie EW, Dobryakova E, Donnat CL, Dukart J, Duncan NW, Durnez J, Eed A, Eickhoff SB, Erhart A, Fontanesi L, Fricke GM, Fu S, Galván A, Gau R, Genon S, Glatard T, Glerean E, Goeman JJ, Golowin SAE, González-García C, Gorgolewski KJ, Grady CL, Green MA, Guassi Moreira JF, Guest O, Hakimi S, Hamilton JP, Hancock R, Handjaras G, Harry BB, Hawco C, Herholz P, Herman G, Heunis S, Hoffstaedter F, Hogeveen J, Holmes S, Hu CP, Huettel SA, Hughes ME, Iacovella V, Iordan AD, Isager PM, Isik AI, Jahn A, Johnson MR, Johnstone T, Joseph MJE, Juliano AC, Kable JW, Kassinopoulos M, Koba C, Kong XZ, Koscik TR, Kucukboyaci NE, Kuhl BA, Kupek S, Laird AR, Lamm C, Langner R, Lauharatanahirun N, Lee H, Lee S, Leemans A, Leo A, Lesage E, Li F, Li MYC, Lim PC, Lintz EN, Liphardt SW, Losecaat Vermeer AB, Love BC, Mack ML, Malpica N, Marins T, Maumet C, McDonald K, McGuire JT, Melero H, Méndez Leal AS, Meyer B, Meyer KN, Mihai G, Mitsis GD, Moll J, Nielson DM, Nilsonne G, Notter MP, Olivetti E, Onicas AI, Papale P, Patil KR, Peelle JE, Pérez A, Pischedda D, Poline JB, Prystauka Y, Ray S, Reuter-Lorenz PA, Reynolds RC, Ricciardi E, Rieck JR, Rodriguez-Thompson AM, Romyn A, Salo T, Samanez-Larkin GR, Sanz-Morales E, Schlichting ML, Schultz DH, Shen Q, Sheridan MA, Silvers JA, Skagerlund K, Smith A, Smith DV, Sokol-Hessner P, Steinkamp SR, Tashjian SM, Thirion B, Thorp JN, Tinghög G, Tisdall L, Tompson SH, Toro-Serey C, Torre Tresols JJ, Tozzi L, Truong V, Turella L, van ’t Veer AE, Verguts T, Vettel JM, Vijayarajah S, Vo K, Wall MB, Weeda WD, Weis S, White DJ, Wisniewski D, Xifra-Porxas A, Yearling EA, Yoon S, Yuan R, Yuen KSL, Zhang L, Zhang X, Zosky JE, Nichols TE, Poldrack RA, Schonberg T. Variability in the analysis of a single neuroimaging dataset by many teams. Nature. 2020 Jun;582(7810):84–88.

21. Ballin M, Ioannidis JPA, Bergman J, Kivipelto M, Nordstrom A, Nordstrom P. Time-Varying Death Risk After SARS-CoV-2-Infection in Swedish Long-Term Care Facilities. medRxiv 2022.03.10.22272097; doi: https://doi.org/10.1101/2022.03.10.22272097

22. Lee RD, Carter LR. Modeling and forecasting US mortality. Journal of the American Statistical Association 1992;87:659–671

23. Booth H, Hyndman RJ, Tickle L, de Jong P. Lee-Carter mortality forecasting: a multi-country comparison of variants and extensions. Demographic Research 2006.15:289– 310.

24. Bergeron-Boucher M-P, Kjærgaard S. Mortality forecasting at age 65 and above: an age-specific evaluation of the Lee-Carter model, Scandinavian Actuarial Journal, 2022;1:64–79

25. https://www.cdc.gov/nchs/data/nhis/earlyrelease/insur201905.pdf, last accessed August 31, 2022.

26. Woolf SH. Progress in achieving health equity requires attention to root causes. Health Aff (Millwood). 2017 Jun 1;36(6):984–991.

27. Sommers BD, Gunja MZ, Finegold K, Musco T. Changes in Self-reported Insurance Coverage, Access to Care, and Health Under the Affordable Care Act. JAMA. 2015 Jul 28;314(4):366–74

28. Flegal KM, Carroll MD, Kit BK, Ogden CL. Prevalence of obesity and trends in the distribution of body mass index among US adults, 1999-2010. JAMA. 2012 Feb 1;307(5):491–7.

29. Kuehn BM. Massive costs of the US opioid epidemic in lives and dollars. JAMA. 2021 May 25;325(20):2040.

30. Bauchner H, Rivara FP, Bonow RO, Bressler NM, Disis MLN, Heckers S, Josephson SA, Kibbe MR, Piccirillo JF, Redberg RF, Rhee JS, Robinson JK. Death by gun violence-a public health crisis. JAMA. 2017 Nov 14;318(18):1763–1764.

31. Health systems in transition series, in: https://eurohealthobservatory.who.int/publications/health-systems-reviews?publicationtypes=e8000866-0752-4d04-a883-a29d758e3413&publicationtypes-hidden=true, last accessed August 31, 2022.

32. Laliotis I, Ioannidis JPA, Stavropoulou C. Total and cause-specific mortality before and after the onset of the Greek economic crisis: an interrupted time-series analysis. Lancet Public Health. 2016 Dec;1(2):e56–e65.

33. Akhtar-Danesh N, Baumann A, Crea-Arsenio M, Antonipillai V. COVID-19 excess mortality among long-term care residents in Ontario, Canada. PLoS One. 2022 Jan 20;17(1):e0262807.

34. August M. Socialize, De-commodify and de-financialize long-term care. Healthc Pap. 2021 Sep;20(1):34–39.

35. Ballin M, Bergman J, Kivipelto M, Nordström A, Nordström P. Excess mortality after COVID-19 in Swedish long-term care facilities. J Am Med Dir Assoc. 2021 Aug;22(8):1574–1580.e8.

36. Vestergaard LS, Nielsen J, Richter L, Schmid D, Bustos N, Braeye T, Denissov G, Veideman T, Luomala O, Möttönen T, Fouillet A, Caserio-Schönemann C, an der Heiden M, Uphoff H, Lytras T, Gkolfinopoulou K, Paldy A, Domegan L, O’Donnell J, de’ Donato F, Noccioli F, Hoffmann P, Velez T, England K, van Asten L, White RA, Tønnessen R, P da , Silva S, P Rodrigues A, Larrauri A, Delgado-Sanz C, Farah A, Galanis I, Junker C, Perisa D, Sinnathamby M, Andrews N, O’Doherty M, Marquess DFP, Kennedy S, Olsen SJ, Pebody R , ECDC Public Health Emergency Team for COVID-19, Krause TG, Mølbak K. Excess all-cause mortality during the COVID-19 pandemic in Europe–preliminary pooled estimates from the EuroMOMO network, March to April 2020. Eurosurveillance. 2020 Jul 2;25(26):2001214.

37. Nielsen J, Mazick A, Andrews N, Detsis M, Fenech TM, Flores VM, Foulliet A, Gergonne B, Green HK, Junker C, Nunes B, O’donnell J, Oza A, Paldy A, Pebody R, Reynolds A, Sideroglou T, Snijders BE, Simon-Soria F, Uphoff H, Van Asten L, Virtanen MJ, Wuillaume F, Mølbak K. Pooling European all-cause mortality: methodology and findings for the seasons 2008/2009 to 2010/2011. Epidemiol Infect. 2013;141(9):1996–2010.

38. Portugal L. Mortality and Excess Mortality: Improving FluMOMO. Journal of Environmental and Public Health, 2021, 1–8.

39. Kjærgaard S, Ergemen YE, Bergeron-Boucher MP, Oeppen J, Kallestrup-Lamb M. Longevity forecasting by socio-economic groups using compositional data analysis. Journal of the Royal Statistical Society Series A 2020;183:1167–1187.

40. Ioannidis JP. The end of the COVID-19 pandemic. Eur J Clin Invest 2022;53:e13782.

41. Stoeldraijer L, van Duin C, van Wissen L, Janssen F. Impact of different mortality forecasting methods and explicit assumptions on projected future life expectancy: the case of the Netherlands. Demographic Research 2013;29:323–354.

42. Human mortality database, short-term mortality fluctuations. In: https://www.mortality.org/Data/STMF, last accessed August 20, 2022.

43. Wilmoth JR, Andreev K, Jdanov D, Glei DA, Boe C, Bubenheim M, Philipov D, Shkolnikov V, Vachon P.. Methods protocol for the human mortality database. University of California, Berkeley, and Max Planck Institute for Demographic Research, Rostock. URL: http://mortality. org [version 31/05/2007], 9, pp.10–11.

44. Jdanov DA, Jasilionis D, Shkolnikov VM and Barbieri M. Human mortality database. Encyclopedia of gerontology and population aging/editors Danan Gu, Matthew E. Dupre. Cham: Springer International Publishing, 2020.

